# Overall Health Effects of mRNA COVID-19 Vaccines in Children and Adolescents A Systematic Review and Meta-Analysis

**DOI:** 10.1101/2023.12.07.23298573

**Authors:** Stine S. Hoffmann, Sebastian Nielsen, Sanne M. Thysen, Ram Duriseti, Christine S. Benn

## Abstract

**Importance:** Phase 3 randomized controlled trials (RCTs) of mRNA COVID-19 vaccines in children and adolescents showed efficacy in preventing COVID-19 infections. Vaccines may have non-specific effects.

**Objective:** Conduct a systematic review and meta-analysis of the phase 3 trials to assess overall and non-specific health effects of the mRNA COVID-19 vaccines.

**Data Sources:** PubMed, Embase, Clinical Trials, Web of Science, and regulatory websites were searched for RCTs of mRNA vaccines. The latest trial data was included.

**Study Selection:** All RCTs conducted with mRNA vaccines BNT162b2 and mRNA-1237 in children and adolescents below 18 years, with placebo, adjuvant, or other vaccines as controls. 1199 studies were screened; six were included in the analysis.

**Data Extraction and Synthesis:** Data on serious adverse events (“SAEs”) and severe adverse events (“Severe AEs”) as well as organ-specific diseases was extracted following the PRISMA reporting guideline, with a focus on non-specific infectious events. Risk Ratios (RRs) comparing vaccine vs placebo were calculated for each vaccine and combined in Mantel-Haenszel estimates.

**Main Outcomes and Measures:** The primary outcomes were SAEs: overall, non-accident SAEs, and infectious SAEs, respectively. Secondary outcomes were Severe AEs and lower respiratory tract infection (LRTI) including RSV.

**Results:** The analyses included 25,549 individuals (17,538 received mRNA; 8,011 received placebo). The risk of SAEs was similar for vaccine and placebo recipients. Both mRNA vaccines were associated with increased risk of severe AEs in older children. In a combined analysis, the RR was 3.77 (1.56-9.13[0.4% vs 0.1% in vaccine vs placebo recipients]) in above 5 year-olds, and 0.82 (0.53-1.29)[0.8% vs 0.9%])in younger children, who received a lower dose of vaccine (p=0.003 for same effect in older and younger children). In the younger children, mRNA vaccines were associated with higher risk of LRTI (RR=2.80 (1.32-5.94)[0.6% vs 0.3%]) including a higher risk of RSV infections (RR=2.78 (1.09-7.06)[0.4% vs 0.2%]).

**Conclusions and Relevance:** mRNA vaccines did not increase the risk of SAEs but were associated with an increased risk of severe AEs in older children, and an increased risk of LRTI, including RSV, in the young. Further research into the overall and non-specific health effects of mRNA vaccines is warranted.

## INTRODUCTION

COVID-19 vaccines were developed and implemented at an unprecedented speed. As of September 2023, 11 COVID-19 vaccines had received licensure or emergency use authorization (EUA). All COVID-19 vaccines were tested in phase 3 randomized clinical trials (RCTs) in adults and showed efficacy in preventing COVID-19 infections. Subsequent phase 3 trials included progressively younger populations. The most administered vaccine types in children and adolescents are the mRNA vaccines (Pfizer/BioNTech (BNT162B2) and Moderna (mRNA-1273)). Both have received full approval or emergency/conditional authorization in the United States and the European Union children from 6 months of age.

Children have biological differences from adults that gradually diminish with age.^1^ Most children with a COVID-19 infection are asymptomatic or have influenza-like symptoms.^2,3^ Given the age-related differences in COVID-19 morbidity, COVID-19 mRNA vaccine benefit-risk assessments may differ in children vs. adults.

Safety data collected during vaccine trials are reported as either “solicited” adverse reactions, i.e., events related to reactogenicity, or as “unsolicited” adverse events (AEs) that are spontaneously reported unexpected events. AEs are divided by severity. Serious AEs (SAEs) are outcomes leading to death, life-threatening events, hospitalization, or disability/permanent damage, also defined as Grade 4 and/or 5 AEs.^4^ Severe AEs are medically significant but not immediately life-threatening events, also known as Grade 3 AEs. In vaccine trials, Severe AEs are usually collected up to 1 month after vaccination, whereas information on SAEs, (all-cause deaths, and hospitalizations) and AEs of special interest (AESIs) should be observed in all study participants for at least 6 months after the last vaccination dose.^5^

Since the implementation of COVID-19 vaccines in children, several AEs have been detected in post-authorization safety monitoring.^6–10^ These include encephalitis,^11^ nephritis,^12^ neurological complications,^13^ and myocarditis.^14^

Vaccines may have non-specific (off-target) effects, i.e., an ability to affect the risk of other diseases by either reducing or increasing the susceptibility to other infections. In general, live vaccines reduce susceptibility to a broad range of other infections, whereas non-live vaccines may increase susceptibility to other infections.^15^ The existence of non-specific effects was not taken into consideration when the current framework for testing vaccine efficacy and safety was established.^16^

In this review, as previously performed for the adults’ phase 2-3-trials,^17^ the available phase 3 trial reports were used to examine the overall health effect of COVID-19 mRNA vaccines in children, with a particular focus on non-COVID-19 infectious disease outcomes.

## METHODS

The systematic review was conducted under the Preferred Reporting Items for Systematic Reviews and Meta-analyses (PRISMA) reporting guideline and registered in PROSPERO before initiating the search.^18,19^

### Search strategy

Randomized controlled trials (RCTs) with participants under 18 years old meeting the following criteria were included: phase 2/3 studies of randomized placebo-controlled trials of COVID-19 mRNA vaccines published in a peer-reviewed journal or in the process of applying for regulatory approval.

### Search for published literature

In March 2023, we searched PubMed, Embase, Clinical Trials, and Web of Science for RCTs of mRNA vaccines (Search string in Supplementary Material). All identified studies were exported into Covidence. The search was rerun in September 2023 to ensure that the review was up to date.

All RCTs with original data, comparing Pfizer and Moderna mRNA vaccines as interventions, with placebo, adjuvant, or other vaccines as controls. After removing duplicates, two review authors (SH and CSB) independently assessed all records.

### Search for regulatory reports

Next, trial data was found on *The United States Food and Drug Administration* (FDA) and *The European Medicines Agency* (EMA) websites. For each study, we obtained the newest vaccine trial data published by the FDA and EMA matching on clinical trial number, local trial number, and number of participants in each age group.^20–27^

### Data extraction

Relevant data was extracted from the most comprehensive report. The following data were extracted (where possible): study characteristics (including author, publication year, local Pfizer/Moderna trial number and Clinical Trial number), age and sex of participants, enrollment period and location, trial phase, type of vaccine and placebo, number of vaccines, antigen dose per injection, and health outcome data.

### Health outcome data

The three co-primary outcomes were SAEs: (a) overall, b) excluding accidents, and c) infectious). Additional outcomes were Infectious disease deaths; Infectious disease hospitalization; Severe AEs (overall and infectious); Adverse events of special interest (AESIs), and other health outcomes where reporting and numbers allow for meaningful analysis, with a focus on non-COVID infectious diseases.

For efficacy estimates and the safety outcomes SAEs, AESIs, and death, participants had been followed for the full duration of the study (Table 1). The less severe outcomes, Severe AEs and other health outcomes (defined by System Organ Class (SOC)), were only registered for one month after each vaccine dose. The trials in the younger children reported specific data on respiratory infections, including pneumonia, and pathogen-specific disease such as RSV. As non-specific effects of vaccines have been shown to be most pronounced for lower respiratory infections (LRTIs),^28^ we analyzed the effect of the vaccines on LRTI, which we defined as pneumonia, lower tract infection, and/or RSV infection, and we furthermore subdivided LRTI into RSV infection and other LRTI. “Upper respiratory tract infections” (URTI), defined as upper respiratory infections (URIs), viral URI, and croup, were also analyzed.

**Table 1.**
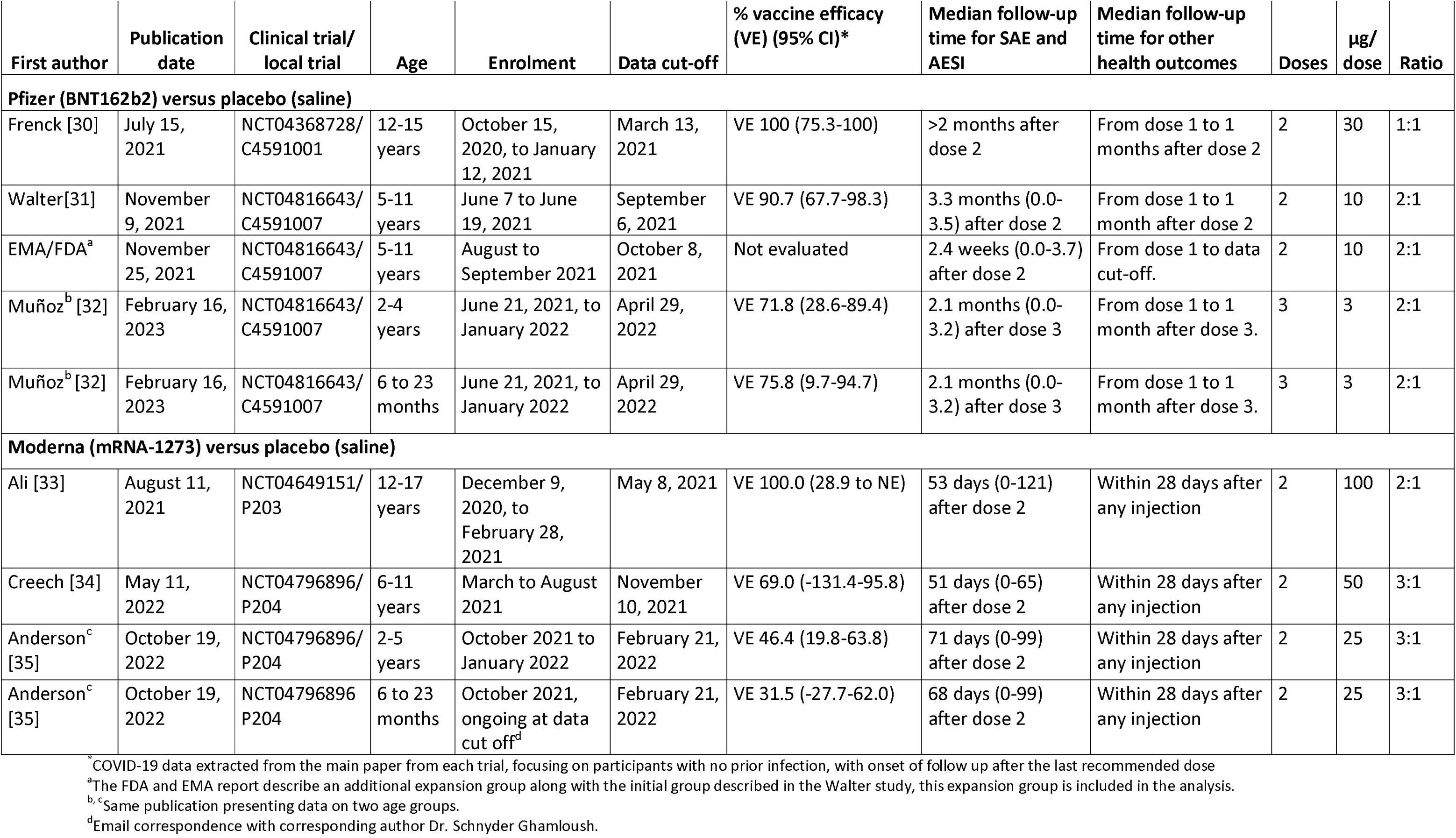
Background information on the included phase 3 studies of COVID-19 vaccination of children.

AEs could be registered as both Severe AEs and SAEs according to event definition in each trial’s supplementary material, AESIs could also be SAEs, and respiratory infections could also be registered as Severe AEs or SAEs thereby preventing composite outcome analysis.

Data on vaccine efficacy against COVID-19 was extracted from each trial’s main publications plus supplementary material. We chose the local trial definition of COVID-19 infection.

All health outcomes and more detail on data extraction are presented in Supplementary Tables 1 and 2. Cochrane Risk of Bias tool (RoB) for RCTs was used to assess the quality of the included publications (Supplementary Table 3).

### Data analysis

All analyses were done by age group (6-23 months, 2-4(5)-years, 5(6)-11 years, 12-15(17) years) and vaccine type. There was insufficient information to analyze data by sex.

The absolute risks for vaccine and placebo groups are presented with the number of events as X% (n). Risk ratios (RRs) with 95% confidence intervals were calculated using the *metan* command in Stata BE version 18 providing Mantel-Haenszel (MH) estimates. However, for age groups with zero events of a certain outcome in one of the groups, RRs were calculated using the Peto OR method, which provides a better approximation than MH, as the MH method applies a continuity correction of 0.5 by default.^29^ In such cases ORs are presented in the tables (marked with *). The *metan* command was also used to calculate combined MH fixed effects estimates, as recommended over random effects estimates in the case of rare events^29^ and to draw Forest plots and conduct test of homogeneity of estimates across strata. All combined estimates were calculated stratified for the variables by which they were combined (age group and/or vaccine type).

We compared the Number Needed to Treat (NNT) separately for the four age groups and overall, for preventing a case of COVID-19 to the risk of different adverse event outcomes. We calculated the NNT as 1/{[control group risk]^RR^ – control group risk}. The corresponding 95% confidence intervals was calculated by replacing the RR with the lower and higher confidence limits.^30^ A positive NNT indicates the number of children that need to be vaccinated to prevent one event (hereafter the number needed to treat to benefit (NNTB)). A negative NNT implies that the vaccine is more likely to harm than benefit and the absolute value may also be interpreted as number needed to treat to harm (NNTH).^30^

## RESULTS

The main search for published literature resulted in 1864 studies from databases/registers and 14 references from other sources. After removing duplicates, 1199 records were screened, 1135 were excluded, and 64 full texts were assessed. Of these, six studies were included in the analysis.^31–36^ (Supplementary Figure 1). For each of the two mRNA COVID-19 vaccines, three main publications were published according to age groups: 12-15 years,^31^ 5-11 years,^32^ and 6 months to 4 years for Pfizer (presenting safety data for the 6 to 23 months and 2-4 year old groups separately),^33^ and for Moderna 12-17 years,^34^ 6-11 years,^35^ and 6 months to 5 years (presenting safety data for the 6-23 months and 2-5 year old groups separately).^36^ Table 1 presents background information on the included phase 3 studies.

### Primary outcome: SAEs (three definitions: a) overall, b) non-accidents, and c) infectious)

There were no reported deaths in any of the trials. There was one SAE due to COVID-19, in the placebo group of 5-11-year-old children. The combined RR for overall SAEs for mRNA vaccine vs placebo recipients was 1.12 (0.74-1.70) (Table 2). For the Pfizer vaccine, the RR for SAEs tended to be highest in the older children (3.49 (0.73-16.78[0.6% vs 0.2% in vaccine vs placebo recipients]) and declined with lower age. For Moderna vaccines, the highest RR was seen among the youngest population (5.02 (0.66-37.90[0.9% vs 0.2%])), but overall numbers were small (Figure 1, Supplementary tables 4a+b). Splitting data by above and below 5(6) years of age, the combined estimates were 1.84 (0.81-4.20[0.2% vs 0.1%]) for the oldest children and 0.93 (0.57-1.51[0.7% vs 0.8%]) for the youngest (p for homogeneity of effects=0.20)(Figure 1). The results were similar when excluding accidents (1.11 (0.72-1.72)) and when focusing on infectious SAEs (1.08 (0.41-2.83)) (Table 2).

**Figure 1.**
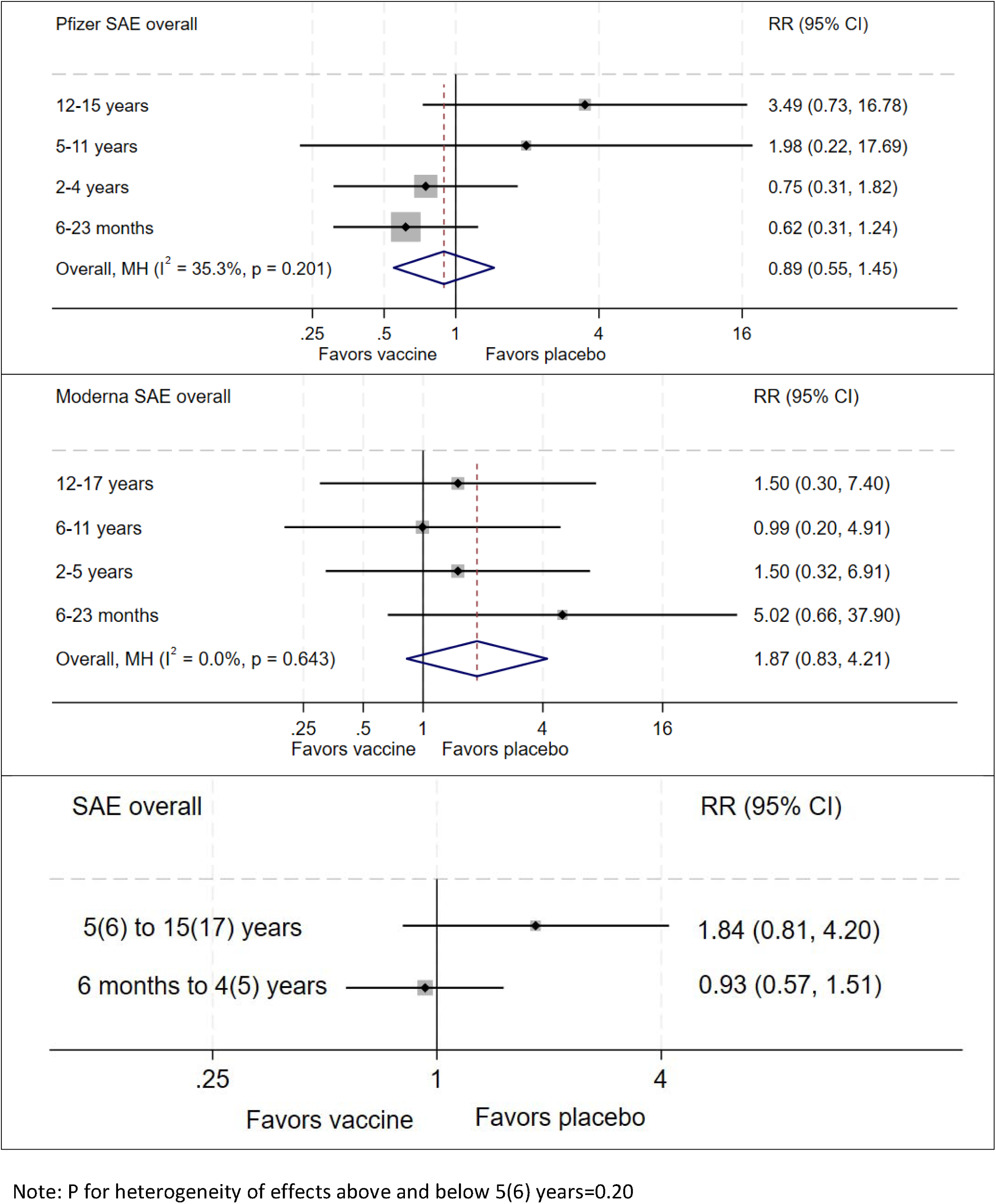
The risk of Overall SAEs in the mRNA phase 3 trials. Analysis done by vaccine type as well as combined, comparing children by age group, and children older and younger than 5(6) years of age, presenting RR (95% CI) and p for homogeneity of effects across age groups.

**Table 2.**
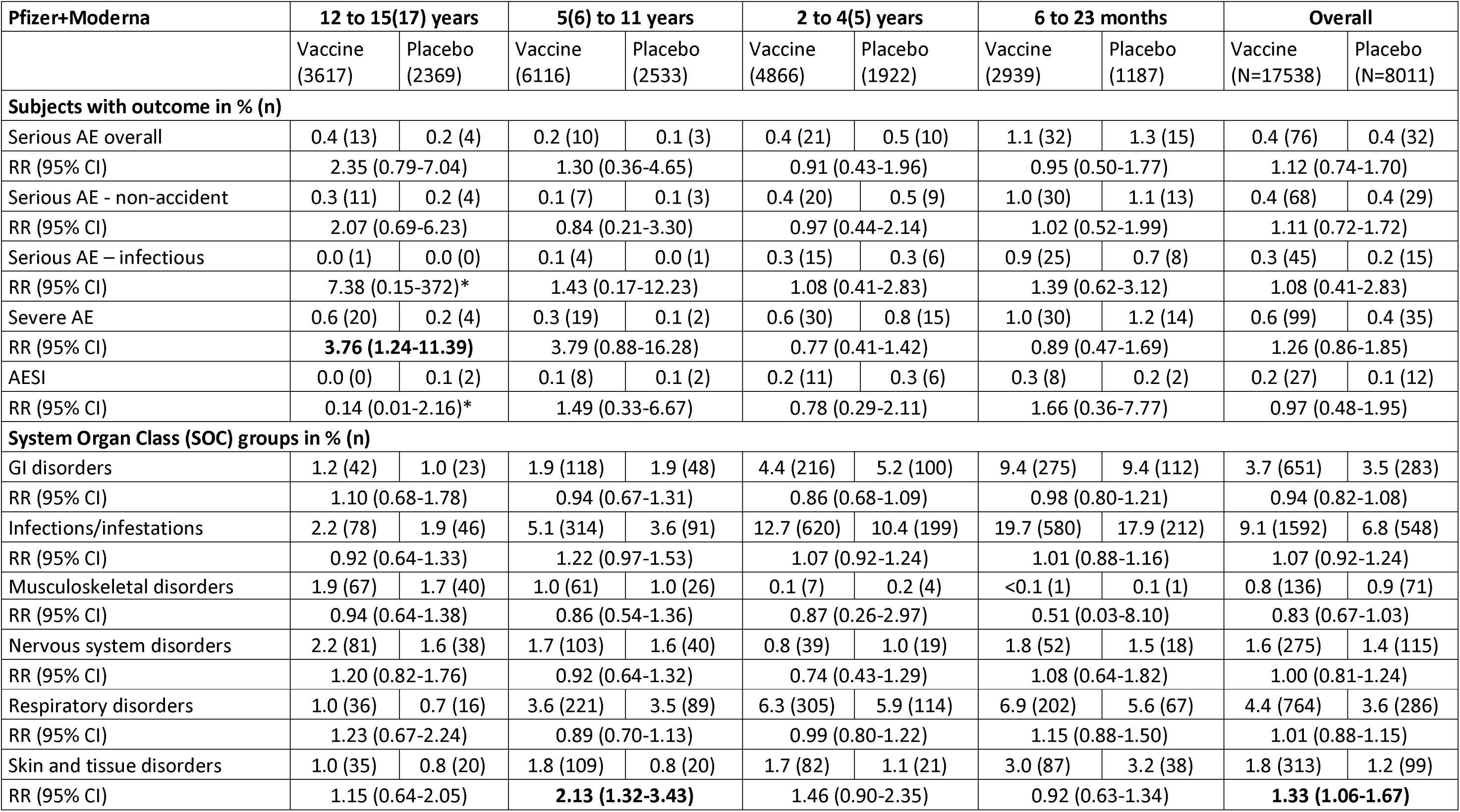
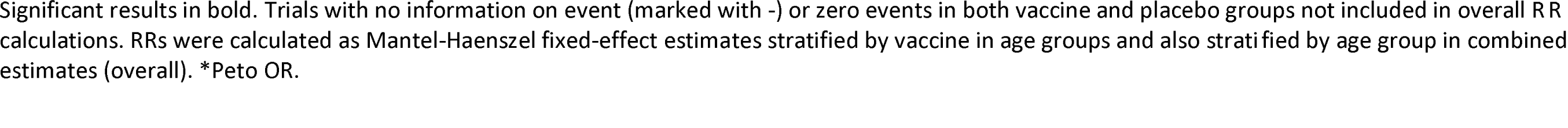
The risk of serious adverse events, severe adverse events, AESI and health outcomes by System Organ Class for children included in the mRNA phase 3 trials, combining Pfizer and Moderna versus placebo.

| Pfizer+Moderna | 12 to 15(17) years |  | 5(6) to 11 years |  | 2 to 4(5) years |  | 6 to 23 months |  | Overall |  |
| --- | --- | --- | --- | --- | --- | --- | --- | --- | --- | --- |
|  | Vaccine<br>(3617) | Placebo<br>(2369) | Vaccine<br>(6116) | Placebo<br>(2533) | Vaccine<br>(4866) | Placebo<br>(1922) | Vaccine<br>(2939) | Placebo<br>(1187) | Vaccine<br>(N=17538) | Placebo<br>(N=8011) |
| Subjects with outcome in % (n) |  |  |  |  |  |  |  |  |  |  |
| Serious AE overall | 0.4 (13) | 0.2 (4) | 0.2 (10) | 0.1 (3) | 0.4 (21) | 0.5 (10) | 1.1 (32) | 1.3 (15) | 0.4 (76) | 0.4 (32) |
| RR (95% CI) | 2.35 (0.79-7.04) |  | 1.30 (0.36-4.65) |  | 0.91 (0.43-1.96) |  | 0.95 (0.50-1.77) |  | 1.12 (0.74-1.70) |  |
| Serious AE - non-accident | 0.3 (11) | 0.2 (4) | 0.1 (7) | 0.1 (3) | 0.4 (20) | 0.5 (9) | 1.0 (30) | 1.1 (13) | 0.4 (68) | 0.4 (29) |
| RR (95% CI) | 2.07 (0.69-6.23) |  | 0.84 (0.21-3.30) |  | 0.97 (0.44-2.14) |  | 1.02 (0.52-1.99) |  | 1.11 (0.72-1.72) |  |
| Serious AE – infectious | 0.0 (1) | 0.0 (0) | 0.1 (4) | 0.0 (1) | 0.3 (15) | 0.3 (6) | 0.9 (25) | 0.7 (8) | 0.3 (45) | 0.2 (15) |
| RR (95% CI) | 7.38 (0.15-372)* |  | 1.43 (0.17-12.23) |  | 1.08 (0.41-2.83) |  | 1.39 (0.62-3.12) |  | 1.08 (0.41-2.83) |  |
| Severe AE | 0.6 (20) | 0.2 (4) | 0.3 (19) | 0.1 (2) | 0.6 (30) | 0.8 (15) | 1.0 (30) | 1.2 (14) | 0.6 (99) | 0.4 (35) |
| RR (95% CI) | 3.76 (1.24-11.39) |  | 3.79 (0.88-16.28) |  | 0.77 (0.41-1.42) |  | 0.89 (0.47-1.69) |  | 1.26 (0.86-1.85) |  |
| AESI | 0.0 (0) | 0.1 (2) | 0.1 (8) | 0.1 (2) | 0.2 (11) | 0.3 (6) | 0.3 (8) | 0.2 (2) | 0.2 (27) | 0.1 (12) |
| RR (95% CI) | 0.14 (0.01-2.16)* |  | 1.49 (0.33-6.67) |  | 0.78 (0.29-2.11) |  | 1.66 (0.36-7.77) |  | 0.97 (0.48-1.95) |  |
| System Organ Class (SOC) groups in % (n) |  |  |  |  |  |  |  |  |  |  |
| GI disorders | 1.2 (42) | 1.0 (23) | 1.9 (118) | 1.9 (48) | 4.4 (216) | 5.2 (100) | 9.4 (275) | 9.4 (112) | 3.7 (651) | 3.5 (283) |
| RR (95% CI) | 1.10 (0.68-1.78) |  | 0.94 (0.67-1.31) |  | 0.86 (0.68-1.09) |  | 0.98 (0.80-1.21) |  | 0.94 (0.82-1.08) |  |
| Infections/infestations | 2.2 (78) | 1.9 (46) | 5.1 (314) | 3.6 (91) | 12.7 (620) | 10.4 (199) | 19.7 (580) | 17.9 (212) | 9.1 (1592) | 6.8 (548) |
| RR (95% CI) | 0.92 (0.64-1.33) |  | 1.22 (0.97-1.53) |  | 1.07 (0.92-1.24) |  | 1.01 (0.88-1.16) |  | 1.07 (0.92-1.24) |  |
| Musculoskeletal disorders | 1.9 (67) | 1.7 (40) | 1.0 (61) | 1.0 (26) | 0.1 (7) | 0.2 (4) | <0.1 (1) | 0.1 (1) | 0.8 (136) | 0.9 (71) |
| RR (95% CI) | 0.94 (0.64-1.38) |  | 0.86 (0.54-1.36) |  | 0.87 (0.26-2.97) |  | 0.51 (0.03-8.10) |  | 0.83 (0.67-1.03) |  |
| Nervous system disorders | 2.2 (81) | 1.6 (38) | 1.7 (103) | 1.6 (40) | 0.8 (39) | 1.0 (19) | 1.8 (52) | 1.5 (18) | 1.6 (275) | 1.4 (115) |
| RR (95% CI) | 1.20 (0.82-1.76) |  | 0.92 (0.64-1.32) |  | 0.74 (0.43-1.29) |  | 1.08 (0.64-1.82) |  | 1.00 (0.81-1.24) |  |
| Respiratory disorders | 1.0 (36) | 0.7 (16) | 3.6 (221) | 3.5 (89) | 6.3 (305) | 5.9 (114) | 6.9 (202) | 5.6 (67) | 4.4 (764) | 3.6 (286) |
| RR (95% CI) | 1.23 (0.67-2.24) |  | 0.89 (0.70-1.13) |  | 0.99 (0.80-1.22) |  | 1.15 (0.88-1.50) |  | 1.01 (0.88-1.15) |  |
| Skin and tissue disorders | 1.0 (35) | 0.8 (20) | 1.8 (109) | 0.8 (20) | 1.7 (82) | 1.1 (21) | 3.0 (87) | 3.2 (38) | 1.8 (313) | 1.2 (99) |
| RR (95% CI) | 1.15 (0.64-2.05) |  | 2.13 (1.32-3.43) |  | 1.46 (0.90-2.35) |  | 0.92 (0.63-1.34) |  | 1.33 (1.06-1.67) |  |

Significant results in bold. Trials with no information on event (marked with -) or zero events in both vaccine and placebo groups not included in overall RR calculations. RRs were calculated as Mantel-Haenszel fixed-effect estimates stratified by vaccine in age groups and also stratified by age group in combined estimates (overall). \*Peto OR.

### Severe AEs

Severe AEs were not reported as narratives, so it was not possible to distinguish between overall and infectious Severe AEs. For both vaccines there was higher risk of Severe AEs in vaccine vs placebo recipients in the two oldest populations, but not in the two youngest populations. For the 12-15(17)-year-olds, the combined RR was 3.76 (1.24-11.39[0.6% vs 0.2%])). In the 5(6)-12-year-olds, the same tendency was observed (3.79 (0.88-16.28[0.3% vs 0.1%])). The overall risk of Severe AEs for younger children was lower in vaccine recipients (Table 2). Splitting data by above and below 5(6) years of age, the estimates were significantly different for the oldest (3.77 (1.56-9.13)[0.4% vs 0.1%]) and the youngest children (0.82 (0.53-1.29)[0.8% vs 0.9%]), p for homogeneity of effect=0.003 (Figure 2).

**Figure 2.**
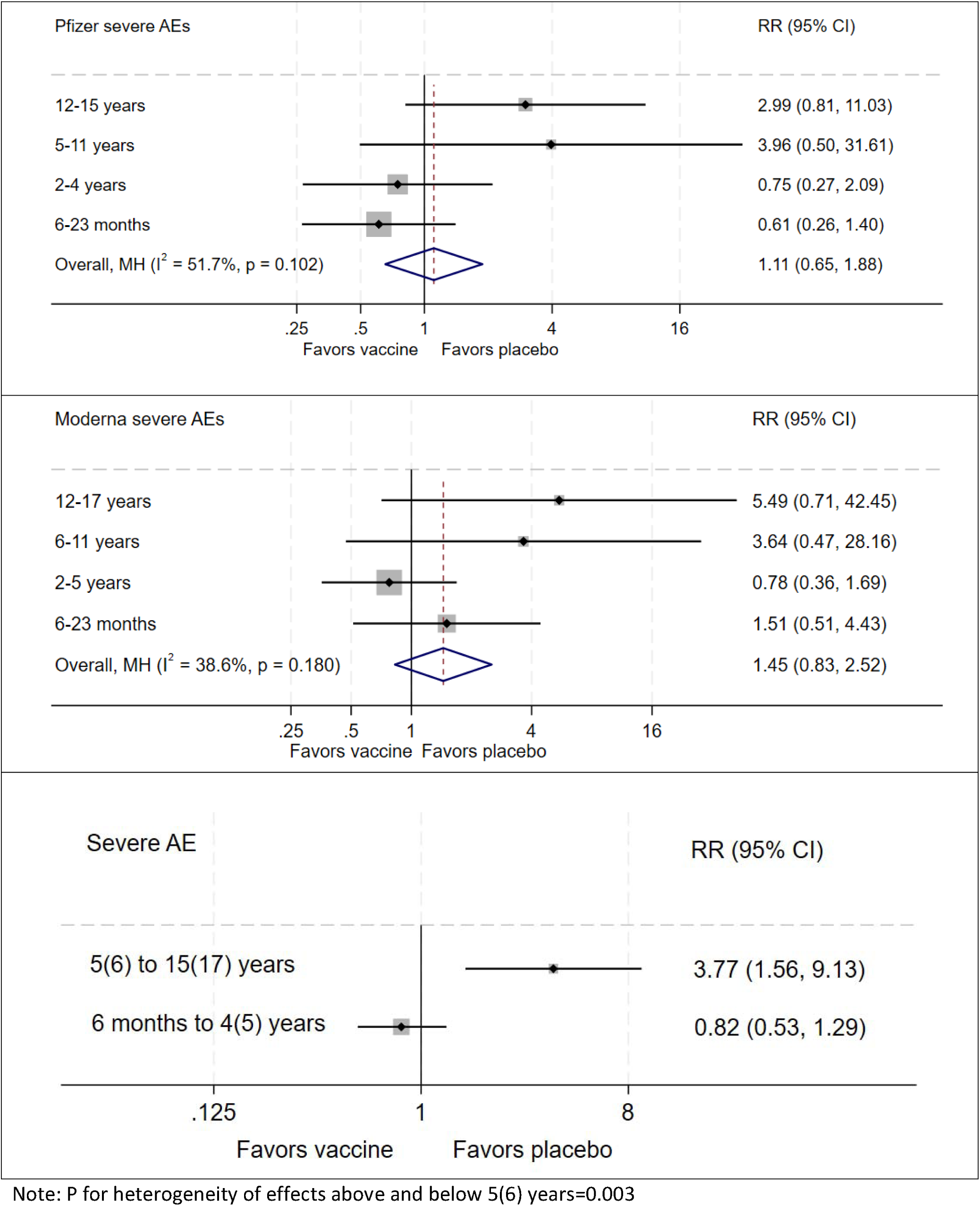
The risk of severe AEs in the mRNA phase 3 trials. Analysis done by vaccine type as well as combined, comparing children by age group, and children older and younger than 5(6) years of age, presenting RR (95% CI) and p for homogeneity of effects across age groups.

### AESIs

Numbers of events per age group were lower than for SAEs and Severe AEs (Supplementary tables 4a+b). The combined RR was 0.97 (0.48-1.95[0.2% vs 0.1%])(Table 2).

### Respiratory tract infections

The trials only reported respiratory tract infections in the younger age groups. The risk of LRTI was higher in vaccine recipients (2.80 (1.32-5.94)[0.6% vs 0.3%])(Table 3, Figure 3). Moderna reported RSV cases for the three youngest age groups whereas Pfizer only did so for the youngest age groups. Restricting the LRTI to RSV, the risk was also higher in vaccine recipients (2.78 (1.09-7.06)[0.4% vs. 0.2%]). A similar tendency was seen for other LRTI (2.85 (0.80-10.17[0.2% vs. 0.1%])). There was no difference in the risk of URTI (1.01 (0.87-1.16)[4.8% vs. 4.0%])(Table 3, Figure 3). As expected, the risk of COVID-19 infections was significantly lower in vaccine recipients with overall RRs of 0.15 (0.08-0.27[0.5% vs.2.7%]) for Pfizer and 0.55 (0.41-0.74[1.2% vs.2.1%]) for Moderna. For Moderna, only the RR for the 2–5-year-olds in Moderna trials was statistically significant (Table 3).

**Figure 3.**
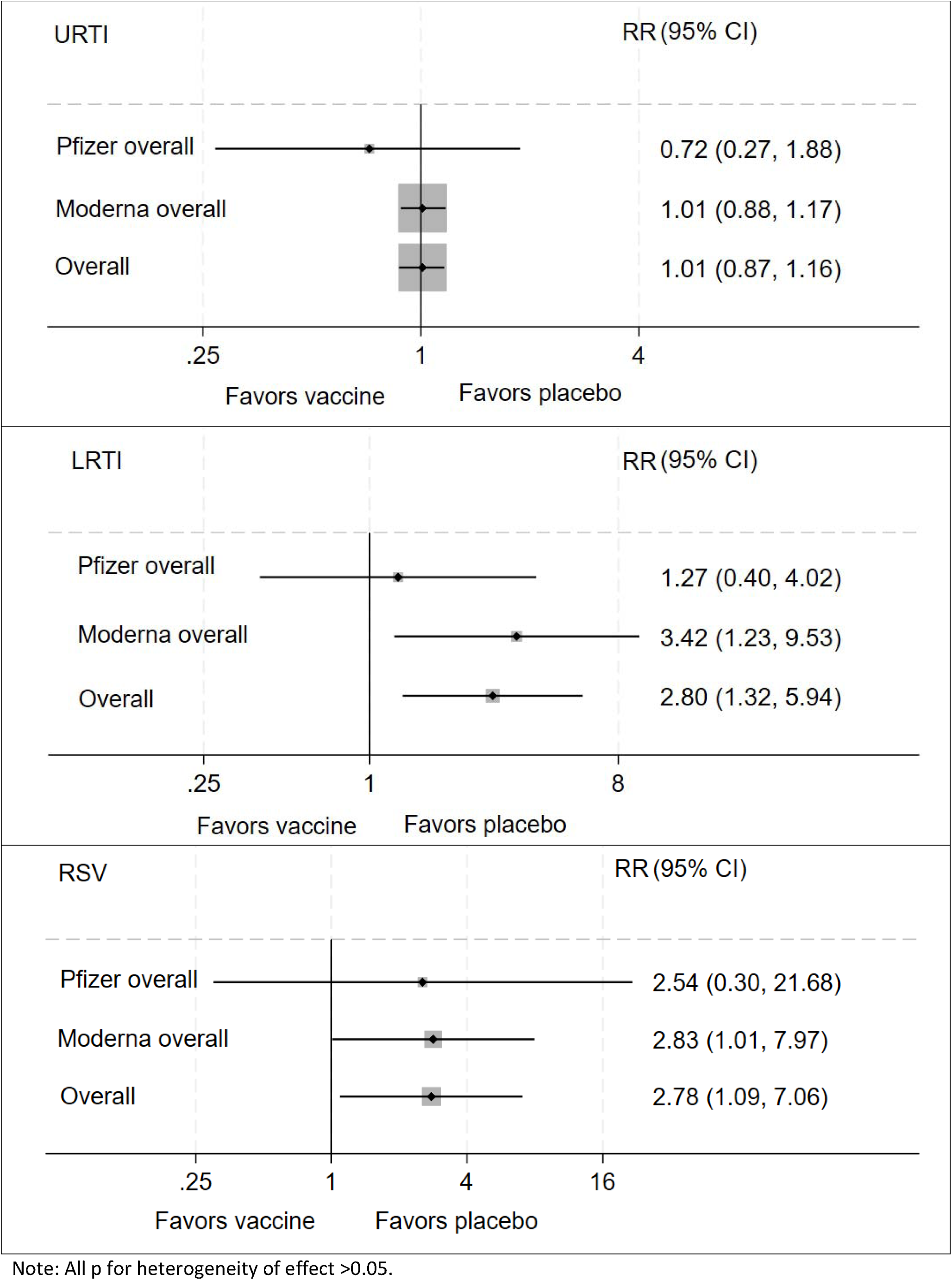
LRTI, RSV and URTI for vaccines versus placebo recipients, according to the age groups where trials had available data. Analysis done by vaccine type as well as combined.

**Table 3.** The risk of respiratory infections for children included in the mRNA phase 3 trials, overall and by vaccine type.

|  | 12 to 15(17) years |  | 5(6) to 11 years |  | 2 to 4(5) years |  | 6 to 23 months |  | Overall |  |
| --- | --- | --- | --- | --- | --- | --- | --- | --- | --- | --- |
|  | Vaccine (3617) | Placebo (2369) | Vaccine (6116) | Placebo (2533) | Vaccine (4866) | Placebo (1922) | Vaccine (2939) | Placebo (1187) | Vaccine | Placebo |
| Subjects with outcome in % (n) |  |  |  |  |  |  |  |  |  |  |
| Upper respiratory infections (URTI) |  |  |  |  |  |  |  |  |  |  |
| Pfizer URTI | - |  | <0.1 (1) | <0.1 (1) | 0.2 (3) | 0.3 (3) | 0.5 (6) | 0.5 (3) | 0.2 (10) | 0.2 (7) |
| Moderna URTI | - | - | 3.9 (116) | 2.5 (25) | 10.4 (316) | 11.1 (112) | 13.2 (232) | 13.8 (81) | 8.5 (664) | 8.4 (218) |
| Overall URTI | - | - | 1.9 (117) | 1.0 (26) | 6.6 (319) | 6.0 (115) | 8.1 (238) | 7.1 (84) | 4.8 (674) | 4.0 (225) |
| Relative Risk (95% CI) | - |  | 1.50 (0.99-2.28) |  | 0.93 (0.76-1.13) |  | 0.96 (0.76-1.21) |  | 1.01 (0.87-1.16) |  |
| Lower respiratory tract infections (LRTI) |  |  |  |  |  |  |  |  |  |  |
| Pfizer LRTI | - | - | - | - | <0.1 (1) | 0.1 (1) | 0.8 (9) | 0.5 (3) | 0.3 (10) | 0.3 (4) |
| Moderna LRTI | - | - | 0.3 (9) | 0 (0) | 0.8 (24) | 0.1 (1) | 1.0 (17) | 0.5 (3) | 0.6 (50) | 0.2 (4) |
| Overall LRTI | - | - | 0.3 (9) | 0 (0) | 0.5 (25) | 0.1 (2) | 0.9 (26) | 0.5 (6) | 0.6 (60) | 0.2 (8) |
| Relative Risk (95% CI) | - |  | 3.80 (0.84-17.2)* |  | 4.56 (1.01-19.60) |  | 1.72 (0.71-4.20) |  | 2.80 (1.32-5.94) |  |
| RSV |  |  |  |  |  |  |  |  |  |  |
| Pfizer RSV | - | - | - | - | - | - | 0.4 (5) | 0.2 (1) | 0.4 (5) | 0.2 (1) |
| Moderna RSV | - | - | 0.3 (9) | 0 (0) | 0.4 (11) | 0.1 (1) | 0.8 (14) | 0.5 (3) | 0.4 (34) | 0.2 (4) |
| Overall RSV | - | - | 0.3 (9) | 0 (0) | 0.4 (11) | <0.1 (1) | 0.6 (19) | 0.3 (4) | 0.4 (39) | 0.2 (5) |
| Relative Risk (95% CI) | - |  | 3.80 (0.84-17.2)* |  | 3.65 (0.47-28.27) |  | 1.78 (0.61-5.20) |  | 2.78 (1.09-7.06) |  |
| Lower respiratory tract infections (LRTI), excluding RSV |  |  |  |  |  |  |  |  |  |  |
| Pfizer LRTI, excl. RSV | - | - | - | - | <0.1 (1) | 0.1 (1) | 0.3 (4) | 0.3 (2) | 0.2 (5) | 0.1 (3) |
| Moderna LRTI, excl. RSV | - | - | 0 (0) | 0 (0) | 0.4 (13) | 0 (0) | 0.2 (3) | 0 (0) | 0.2 (16) | 0 (0) |
| Overall LRTI, excl. RSV | - | - | 0 (0) | 0 (0) | 0.3 (14) | 0.1 (1) | 0.2 (7) | 0.2 (2) | 0.2 (21) | 0.1 (3) |
| Relative Risk (95% CI) | - |  | N/A |  | 2.75 (0.87-8.76)* |  | 1.50 (0.36-6.23)* |  | 2.85 (0.80-10.17) |  |
| COVID-19 infection |  |  |  |  |  |  |  |  |  |  |
| Pfizer COVID-19<sup>a</sup> | 0.0 (0/1005) | 1.6 (16/978) | 0.2 (3/1305) | 2.4 (16/663) | 1.8 (9/498) | 6.4 (13/204) | 1.4 (4/296) | 5.4 (8/147) | 0.5 (16/3104) | 2.7 (53/1992) |
| Moderna COVID-19<sup>b</sup> | 0.0 (0/2163) | 0.4 (4/1073) | 0.4 (3/2644) | 0.1 (3/853) | 2.7 (71/2594) | 5.0 (43/858) | 2.4 (37/1511) | 3.5 (18/513) | 1.2 (111/8912) | 2.1 (68/3297) |
| **Overall COVID-19** | 0.0<br>(0/3168) | 1.0<br>(20/2051) | 0.2<br>(6/3949) | 1.3<br>(19/1516) | 2.6<br>(80/3092) | 5.3 (56/1062) | 2.3<br>(41/1807) | 3.9<br>(26/660) | 1.1<br>(127/12016) | 2.3<br>(121/5289) |
| **Relative Risk (95% CI)** | **0.11 (0.05-0.26)\*** |  | **0.14 (0.05-0.35)** |  | **0.49 (0.35-0.68)** |  | **0.57 (0.35-0.93)** |  | **0.39 (0.30-0.50)** |  |

All data from SOC tables except COVID-19 infections. <sup>a</sup> COVID-19 data extracted from the main paper from each trial (according to trial definition), focusing on participants with no prior infection, with onset of follow up after the last recommended dose. <sup>b</sup> COVID-19 data extracted from the main paper from each trial (COVE trial definition), focusing on participants with no prior infection, with onset of follow up after the last recommended dose
Significant results in bold. Trials with no information on event (marked with -) or zero events in both vaccine and placebo groups not included in overall RR calculations. RRs were calculated as Mantel-Haenszel fixed-effect estimates stratified by vaccine in age groups and also stratified by age group in combined estimates (overall). \*Peto OR.

### Other health outcomes defined by System Organ Class

There were no clear patterns in the presented System Organ Class systems. However, “skin and tissue disorders” had a combined RR of 1.33 (1.06-1.67[1.8% vs. 1.2%])(Table 2).

### Number needed to treat

For COVID-19 infections, the combined NNTB ranged from 40 to 115. For SAEs and Severe AEs, the NNT range included both possibilities of benefit and harm, with a range from -2817 to 2141. For severe AE in the oldest age group, the NNTH was -215. For LRTI in younger children, the overall NNTH was -217 and for RSV it was -635 (Supplementary table 5).

## DISCUSSION

From the RCTs in children aged 6 months to 15(17) years, neither of the mRNA vaccines had a beneficial effect on overall health, assessed as the incidence of SAEs, Severe AEs, RTIs, and other organ-specific diseases. mRNA vaccines were not associated with an increased risk of SAEs, but the risk of Severe AEs was more than 3.5-fold higher in the older children. In younger children, specific information on respiratory infections was provided, and mRNA vaccines were associated with a nearly 3-fold increased risk of LRTI, including a nearly 3-fold increase in the risk of RSV. As expected, the vaccines reduced the risk of COVID-19 infection.

### Strengths and limitations

The review was based on placebo controlled RCTs and represents the best available estimates of the overall health effect of COVID-19 vaccines in children. There is unlikely to be publication bias.

Our analyses have several limitations, which are largely a consequence of limited data availability and low reporting standards.

First, the included trials were designed to collect and analyze safety and efficacy of vaccines, not overall health effects. Only SAEs and AESI were collected for the full duration. All health outcomes ought to be investigated.^16,37^ The occurrence (or absence) of e.g., another respiratory infection several months after randomization may well be the result of the vaccination.^15^ While it is not customary to collect all health outcomes for the full duration of vaccine trials, the results on respiratory infections presented here suggest such data can provide important insights.

Second, vaccine efficacy in the younger populations has primarily been assessed by “immunobridging”, where antibody responses in children receiving the vaccine are compared to antibody responses in older age groups, in which efficacy was demonstrated. Hence, clinical trial populations including children were much smaller than in adults. This led to limitations regarding safety analysis, as also highlighted in the EMA report.^21^

Third, the analysis of unsolicited SAEs, Severe AEs, and AESIs was challenging due to inconsistencies in the reported number of events between the main texts, supplementary data, and trial reports: a serious problem that has been previously addressed.^38,39^ Furthermore, the data was reported with an overlap in case definitions that hampered composite outcome analysis.

Fourth, there were considerable differences between the two vaccine types regarding the incidence of reported events, e.g., “infections and infestations” (incidence 13.5% vs 2.8% for Moderna vs Pfizer vaccinees), “nervous system disorders” (2.2% vs 0.7%), and “respiratory disorders” (5.6% vs. 0.5%). The difference in infectious outcomes could be related to Fall dates for the Moderna trial versus Summer dates for the Pfizer trials, but may also reflect different reporting standards or dosing, complicating combined analyses.

Fifth, data was not reported by sex. Non-specific effects have been shown to be sex-differential, and in several instances, an overall null-effect has been shown to be a result of a positive effect in one sex but a negative effect in another.^40,41^ As it pertains to COVID-19 mRNA vaccines and adults, there is a striking female predominance in severe allergic reactions.^42^ Many journals require data be broken down by biological sex.^43^ It is a serious omission that this is still not done consistently.^16^

### Interpretation

For SAEs and Severe AEs, the oldest populations generally had the highest risk compared with the youngest. The observed risk difference could be an effect of the vaccine dose. The youngest children received a lower dose than the 5-11-year-olds, who received a lower than adolescents (Pfizer 3 vs 10 vs. 30 μg, Moderna 25 vs 50 vs 100 μg). The Pfizer dose for the youngest children is 10 times lower than for the oldest group, whereas the Moderna dose was only 4 times lower, which might help explain the stronger dose-response in Severe AEs and SAEs in the Pfizer trials. During the open-label dose-level finding investigation, children 2 to 5 years of age in the Pfizer trials had a higher frequency and greater severity of adverse reactions to the 10 µg dose compared with the 3 µg dose, which contributed to the selection of the lower dose.^21^

Our finding of a higher rate of LRTI after mRNA vaccination lines with other studies showing negative non-specific effects of non-live vaccines for respiratory infections. Sørup et al found a higher risk of both RSV-related and other non-targeted infections in children after receiving a non-live vaccine compared with the live vaccine against Measles, Mumps, and Rubella.^44,45^ Two small immunological studies, one in adults and one in children, have found mRNA vaccines to be associated with a decrease in cytokine responses to viral stimulants for 6 months.^46,47^ Similar immunological effects have been observed for other non-live vaccines.^48,49^ Taken together, the results suggest that mRNA vaccines may have negative non-specific immunologic effects, increasing the risk of unrelated infections and lasting at least some months.

Regarding the increased overall risk of “skin and tissue disorders”, this apparent association could be a chance finding. Nevertheless, various skin reactions from COVID-19 mRNA-vaccines have previously been described.^50^

### Number needed to treat to benefit or harm

Only COVID-19, SAEs, and AESIs were captured for more than 4 weeks post-vaccination. Thus, comparing with health outcomes captured for 4 weeks only, there would inherently be a higher baseline risk, biasing towards higher NNTB/H. For the 12–15-year-olds receiving either vaccine, the NNTB for preventing one additional case of COVID-19 was 107, and the NNTH for developing one additional Severe AE 440. Thus, there would – very conservatively - be one Severe AE per four prevented COVID-19 cases (440 NNTH/107 NNTB≈4.11). In the youngest age group, per four prevented COVID-19 cases, one child would develop an LRTI (NNTB for COVID-19 60; NNTH for LTRI 276), and for each six prevented COVID-19 cases, one child would get an RSV infection (NNTH for RSV 382). LRTI including RSV infections in infants and small children are usually more severe than COVID-19 infections.^51^

## CONCLUSION

Both mRNA vaccines approved to children were associated with reductions in the risk of COVID-19 and showed no difference in the risk of SAEs. Among the older children, the vaccines were associated with a 3.5-fold higher risk of Severe AEs. Among children younger than 5 years of age, the mRNA vaccines were associated with an almost 3-fold higher risk of LRTI including RSV. Given the low risk of severe COVID-19 infections in children, the RCTs call for a renewed assessment of the value of COVID-19 vaccination of children and adolescents.

## Data Availability

All data produced in the present work are contained in the manuscript

## Acknowledgements

In relation to the study published by Anderson et al^35^, Dr. Schnyder Ghamloush kindly provided information on the enrolment period for part 2 of the study.

## Data sharing

All analyzed data is freely available in the public domain, as per references.

## Transparency statement

All authors affirm that the manuscript is an accurate and transparent presentation of the studies being reported, no important aspects has been omitted, and all relevant data has been published.

## Author contributions

The study was conceptualized by CSB and SMT. SSH wrote the first draft with input from the other authors. SN provided statistical advice. All authors commented and approved the final version.

