## Supplementary Material for "Overall Health Effects of mRNA COVID-19 Vaccines in Children and Adolescents A Systematic Review and Meta-Analysis"

Stine Hoffmann, MD, PhD[1], Sebastian Nielsen, MSc, PhD[1], Sanne M. Thysen, MD, PhD[2], Ram Duriseti, MD, PhD[3], Christine S. Benn, DMSc[1,4]

### Supplementary Material

#### Search String

(COVID-19 OR SARS-CoV-2 OR 2019-nCoV OR coronavirus) AND (5-11 OR 12-15 OR pediatric* OR child* OR infan* OR neonat* OR teen* OR adolescen*) AND (vaccine* OR COVID-19 vaccine OR SARS-CoV-2 vaccine OR BNT162b2 OR Pfizer-BioNTech OR Pfizer OR mRNA-1273 OR Moderna OR Spikevax) AND (randomized controlled trial OR double blind method OR clinical trial OR phase II OR phase III)

**Supplementary Figure 1 PRISMA flowchart of the included studies.**

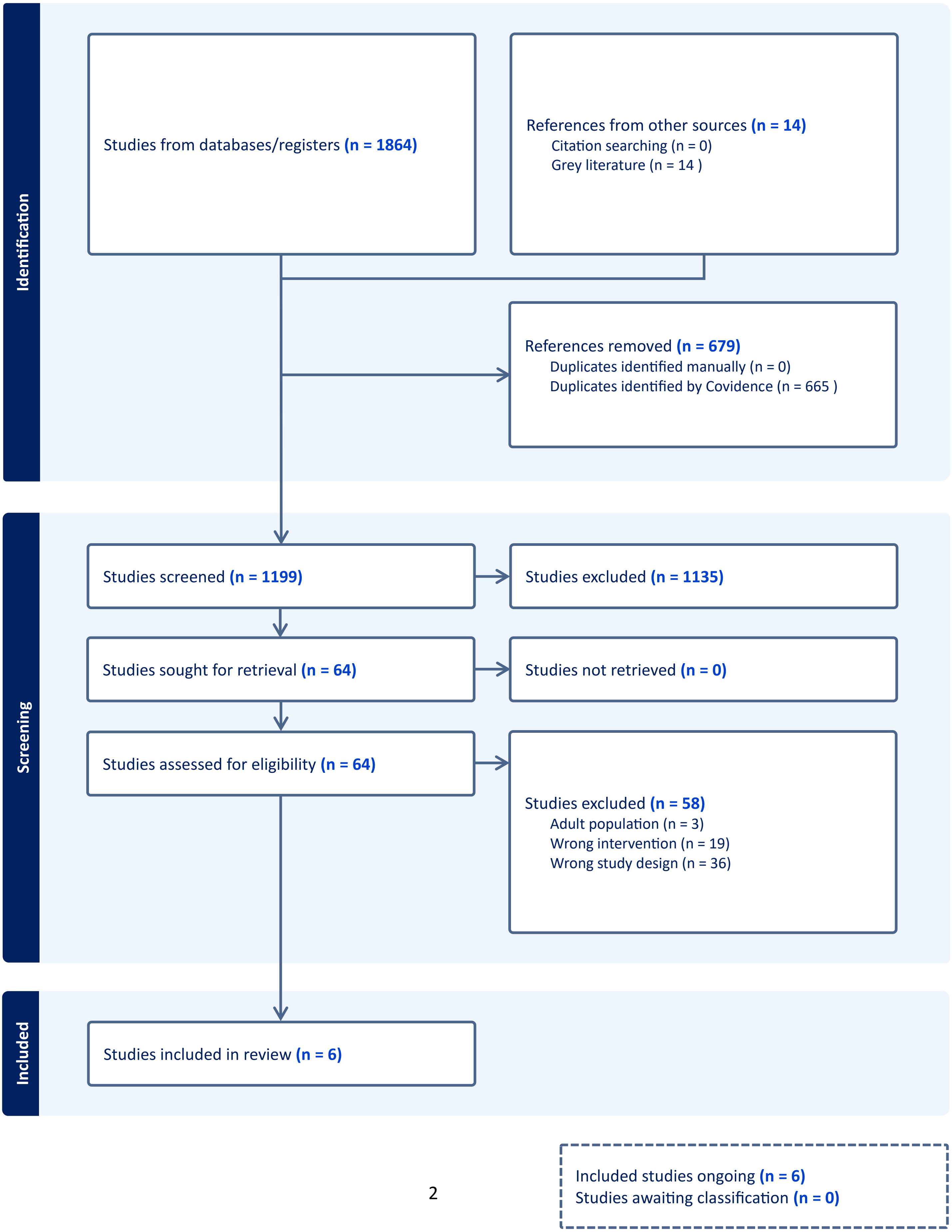

**Supplementary Table 1 a-j.** Description of SAEs in the included studies according to vaccine type and age group. SAE accidents in red, SAE infections in blue. Assessment (related or non-related to vaccine during phase 3) by the trial study investigators are also presented.

We did not distinguish between events judged to be related vs non-related by the study investigators, but merely extracted the total number of events (only very few events were judged as being related). We focused on the follow-up period with onset after the last recommended vaccine dose. In case of more than one table with the same outcome data, we used the publication/report presenting the most recent cut-off date. We only included data with blinded follow-up. In studies reporting data from Part 1 (dose selection with no placebo-receiving control group) and Part 2 (safety and efficacy with chosen dose), we only included data from Part 2.

a) SAE 12-15 years Pfizer[2]

| Treatment group | Preferred term | Time to Onset | Risk Factors, Pertinent Details | Assessment |
|---|---|---|---|---|
| BNT162b2 | depression exacerbation | 7 days after dose 1 | pre-existing anxiety and depression | Non-related |
| BNT162b2 | depression exacerbation | 1 day after dose 2 | pre-existing anxiety and depression | Non-related |
| BNT162b2 | depression exacerbation | 15 days after dose 1 | pre-existing anxiety and depression | Non-related |
| BNT162b2 | Generalized neuralgia, constipation | - | - | Non-related |
| BNT162b2* | Anaphylactoid reaction | 3 days after dose 1 | History of allergy | Related |
| BNT162b2* | Depression | 7 days after dose 1 | - | Non-related |
| BNT162b2 | Pyrexia 40.4 | 2 days after dose 1 | - | Related |
| Placebo | Appendicitis | - | - | Non-related |
| Placebo | Appendicitis | 19 days after dose 2 | - | Non-related |

**2 adolescents originally randomized to the placebo group had SAEs that occurred after they turned 16 years of age during the study and were unblinded to receive BNT162b2, therefore the data are not included in the blinded analyses. These events were also considered as life-threatening: an anaphylactoid reaction reported in 1 participant 3 days after receiving the first dose of BNT162b2 (Dose 3) with a duration of 1 day, considered by the investigator as related to study intervention and leading to study withdrawal; and depression reported in 1 participant 7 days after receiving the first dose of BNT162b2 (Dose 3) reported as ongoing/resolving at the time of the data cut-off date, considered by the investigator as not related to study intervention.*

b) SAE 12-17 years Moderna[3,4]

| Treatment group | Preferred term | Time to Onset | Risk Factors, Pertinent Details | Assessment |
|---|---|---|---|---|
| mRNA-1273 | depression and/or suicidal ideation | 31 days after dose 1 | Depression | Non-related |
| mRNA-1273 | depression and/or suicidal ideation | 30 days after dose 2 | Depression | Non-related |
| mRNA-1273 | depression and/or suicidal ideation | 35 days after dose 2 | - | Non-related |
| mRNA-1273 | Appendicitis | 4 days after dose 1 | | Non-related |
| mRNA-1273 | drug induced-liver injury | 14 days after dose 1 | drug-induced liver injury secondary to trimethoprim/sulfamethoxazole | Non-related |
| mRNA-1273 | surgical repair of pectus excavatum | 31 days after dose 2 | - | Non-related |
| Placebo | Suicide attempt | - | - | Non-related |
| Placebo | Obstructive nephropathy | - | - | Non-related |

c) Additional follow-up – data **not** included in analysis as there is no information on placebo recipients.
*SAEs Captured From Data Cutoff of May 8, 2021, Through Data Cutoff of January 31, 2022, Participants 12 Through 17 Years of Age, P203*

| Treatment Group | Preferred Term | Time to Onset after Most recent dose | Risk Factors/Pertinent Details | Assessment |
|---|---|---|---|---|
| mRNA-1273 | Depression | 46 days after Dose 2 | History of depression, anxiety, post-traumatic stress disorder, substance use | Not related |
| mRNA-1273 | Sunburn | 67 days after Dose 2 | Sunburn with 2nd and 3rd degree burns | Not related |
| mRNA-1273 | Splenic injury; Femur fracture | 68 days after Dose 2 | All-terrain vehicle collision | Not related |
| mRNA-1273 | Clavicle fracture; Concussion; Facial bone fracture | 80 days after Dose 2 | Skateboard accident | Not related |
| mRNA-1273 | Suicidal ideation | 83 days after Dose 2 | History of anxiety, eating disorder, prior suicidal ideation | Not related |
| mRNA-1273 | Suicide ideation | 109 days after Dose 2 | History of anxiety; reported significant domestic stressors around time of SAE | Not related |
| mRNA-1273 | Appendicitis | 182 days after Dose 2 | | Not related |
| mRNA-1273 | Syringomyelia;<br>Meningitis aseptic | 214 days after Dose 2<br>308 days after Dose 2 | History of Chiari I malformation; new symptomatic cervical cord syrinx requiring decompression surgery; developed aseptic meningitis, thought to be related to recent surgery | Not related<br>Not related |
| mRNA-1273 | Suicide attempt | 220 days after Dose 2 | AEs of depression (moderate) and anxiety (mild) 23 days after Dose 2 and was started on antidepressant medication. Reported worsening depression starting 202 days after Dose 2, prior to this SAE. | Not related |
| mRNA-1273 | Major depression | 225 days after Dose 2 | New diagnosis | Not related |
| mRNA-1273 | Suicide attempt | 235 days after Dose 2 | AE of depression (moderate) 208 days after Dose 2 and was started on medication | Not related |
| mRNA-1273 | Suicide ideation | 240 days after Dose 2 | History of depression | Not related |

| | | | | |
|---|---|---|---|---|
| mRNA-1273 | Murine typhus | 246 days after Dose 2 | | Not related |
| mRNA-1273 | Seizure | 285 days after Dose 2 | New diagnosis of idiopathic epilepsy (generalized) | Not related |
| mRNA-1273 | Suicide attempt | 311 days after Dose 2 | AE of depression (mild) 183 days post Dose 2 and was started on medication | Not related |
| mRNA-1273 | Road traffic accident | 68 days after Open-Label Dose 2 | | Not related |

d) SAE 5-11 years Pfizer[5]

Initial + expansion group

| Treatment group | Preferred term | Time to Onset | Risk Factors, Pertinent Details | Assessment |
|---|---|---|---|---|
| BNT162b2 | upper limb fracture after traumatic accident | 45 days after dose 2 | - | Non-related |
| BNT162b2 | infectious arthritis | 15 day after dose 1 | - | Non-related |
| BNT162b2 | foreign body ingestion | 17 days after dose 1 | - | Non-related |
| BNT162b2 | epiphyseal fracture | 20 days after dose 1 | - | Non-related |
| Placebo | Pancreatitis | 4 days after dose 2 | - | Non-related |

e) SAE 6-11 years Moderna[3,4]

| Treatment Group | SAE Preferred Term | Time to Onset After Most Recent Dose | Risk Factors/ Pertinent Details | Assessment |
|---|---|---|---|---|
| mRNA-1273 | Appendicitis | 26 days after Dose 1 | | Not related |
| 50 µg | Appendicitis | 49 days after Dose 2 | | Not related |
| mRNA-1273 | Cellulitis orbital | 2 days after Dose 2 | | Not related |
| mRNA-1273 50 µg | Cellulitis | 16 days after Dose 2 | Cellulitis on elbow in area of mosquito bite, requiring IV antibiotic treatment | Not related |
| mRNA-1273 50 µg | Type I diabetes mellitus | 35 days after Dose 2 | Parent indicated after diagnosis that participant had been having symptoms prior to study entry | Not related |
| mRNA-1273 50 µg | Urosepsis; pyelonephritis | 31 days after Dose 2 | History of tethered spinal cord | Not related |
| Placebo | Affective disorder | 4 days after Dose 2 | History of depression and mood disorder | Not related |
| Placebo | COVID-19 | 25 days after Dose 2 | COVID-19 diagnosis between Dose 1 and Dose 2; presented with “long COVID” symptoms post Dose 2 (tingling/numbness/weakness) | Not related |

f) Additional follow-up:
*SAEs During Additional Follow-Up From Data Cutoff of November 10, 2021, Through Data Cutoff of February 21, 2022, Participants 6 Through 11 Years of Age, Study P204. Both blinded and open-label cross-over. No data on children from placebo who did not cross over to mRNA-1273, hence data* ***not*** *included in SAE analysis.*

| Treatment Group | Preferred Term | Time to Onset/ Most Recent Dose | Risk Factors/Pertinent Details | Assessment |
|---|---|---|---|---|
| mRNA-1273 50 µg | Abdominal pain upper; Cholecystitis | 21 days/ Dose 1 65 days/ Dose 2 | Gallbladder polyps found on HIDA scan | Not related Not related |
| mRNA-1273 50 µg | Epiphyseal fracture | 45 days/ Dose 2 | Distal fracture in the growth plate of the left big toe; occurred while in dance class | Not related |
| mRNA-1273 50 µg | Suicidal ideation; Disruptive mood dysregulation disorder; Oppositional defiant disorder | 68 days/ Dose 2 70 days/ Dose 2 72 days/ Dose 2 | AEs of ADHD and anxiety around the time of event onset | Not related Not related Not related |
| mRNA-1273 50 µg | Febrile neutropenia; Post-procedural complication | 83 days/ Dose 2 130 days/ Dose 2 | After chemotherapy for ependymoma Afebrile neutropenia and anemia after chemotherapy | Not related Not related |
| mRNA-1273 50 µg | Gastrointestinal disorder | 89 days/ Dose 2 | Vomiting and abdominal pain requiring hospitalization | Not related |
| mRNA-1273 50 µg | Asthma | 89 days/ Dose 2 | History of asthma; hospitalized for asthma exacerbation | Not related |
| mRNA-1273 50 µg | Synovitis | 127 days/ Dose 2 | Transient synovitis of bilateral hips | Not related |
| mRNA-1273 50 µg | Neck pain | 128 days/ Dose 2 | Diagnosed with pinched nerve of neck after falling during skiing | Not related |
| mRNA-1273 50 µg | Suicidal ideation | 164 days/ Dose 2 | Participant's guardian reported that participant may have depression/suicidal ideation and participant was referred to | Not related |

| | | | | |
|---|---|---|---|---|
| | | | psychiatrist after evaluation by primary care physician; started on antidepressant | |
| mRNA-1273 50 µg | Kawasaki's disease | 190 days/ Dose 2 | Diagnosis of streptococcal pharyngitis 2 days prior to event; met Kawasaki criteria and treated with IVIG, echocardiogram normal | Not related |
| Placebo-mRNA-1273 50 µg | Ileus | 1 day/ OL Dose 2 | Complex medical history including imperforate anus status post repair and cecostomy, atrial septal defect, gross motor delay, anxiety; hospitalized for abdominal pain after routine enema, abdominal x-ray suggestive of diffuse ileus, passed oral challenge | Related |

g) SAE 2-4 years Pfizer[6]

| Treatment group | Preferred term | Time to Onset | Risk Factors, Pertinent Details | Assessment |
|---|---|---|---|---|
| BNT162b2 | Pyrexia and viral exanthema | 2 days after dose 2 | - | Related |
| BNT162b2 | status epilepticus | 17 days after dose 1 | Miller-Dieker syndrome and seizures | Non-related |
| BNT162b2 | Epilepsy | 47 days after dose 2 (Influvacc 3 days prior) | Febrile convulsions | Non-related |
| BNT162b2 | Febrile convulsions | 20 days after dose 1 | - | Non-related |
| BNT162b2 | Febrile convulsions | 1 day after dose 2 | Neurodevelopmental disorder | Non-related |
| BNT162b2 | Appendicitis | 105 days after dose 2 | - | Non-related |
| BNT162b2 | Appendicitis | 11 days after dose 2 | - | Non-related |
| BNT162b2 | Viral gastroenteritis | 23 days after dose 2 | - | Non-related |
| BNT162b2 | Dehydration/gastroenteritis | - | - | Non-related |
| BNT162b2 | Dehydration | - | - | Non-related |
| BNT162b2 | Upper respiratory tract infection | - | - | Non-related |
| BNT162b2 | Lower respiratory tract infection | - | - | Non-related |
| Placebo | Adenovirus gastroenteritis | - | - | Non-related |
| Placebo | Gastroenteritis | - | - | Non-related |
| Placebo | Rotavirus gastroenteritis | - | - | Non-related |
| Placebo | Foreign body | - | - | Non-related |
| Placebo | Papilledema | - | - | Non-related |
| Placebo | Epilepsy | - | - | Non-related |
| Placebo | Febrile convulsions | - | - | Non-related |
| Placebo | Bronchial hyperreactivity | - | - | Non-related |

h) SAE 2-5 years Moderna[3,4]

| Treatment Group | SAE Preferred Term | Onset in Days Since Last Dose, Last Dose | Risk Factors, Pertinent Details | Assessment |
|---|---|---|---|---|
| mRNA-1273 25 µg | Metapneumovirus infection | 8 days, Dose 1 | Ongoing bronchopulmonary dysplasia, asthma; previous episode of RSV bronchiolitis; hospitalized requiring oxygen | Not related |
| mRNA-1273 25 µg | Rhinovirus infection | 75 days, Dose 2 | Hospitalized for bronchiolitis | Not related |
| mRNA-1273 25 µg | Adenovirus infection | 3 days, Dose 2 | Hospitalized | Not related |
| mRNA-1273 25 µg | Epstein-Barr virus infection | 52 days, Dose 2 | Hospitalized for acute EBV, metabolic acidosis, and dehydration; Diagnosed with mild COVID-19 one month prior; Tested positive for SARS-CoV-2 day before admission; Tested positive for EBV and negative for SARS-CoV-2 during hospitalization | Not related |
| mRNA-1273 25 µg | Pneumonia viral<br>Bronchial hyperreactivity<br>Respiratory distress | 13 days, Dose 1<br>14 days, Dose 1<br>14 days, Dose 1 | None reported; event started as upper respiratory tract infection, left acute otitis media; hospitalized | Not related<br>Not related<br>Not related |
| mRNA-1273 25 µg | Urinary tract infection | 38 days, Dose 2 | Ongoing constipation with fecaloma/colon impaction | Not related |
| mRNA-1273 25 µg | Humerus fracture | 53 days, Dose 2 | Fell while being chased by a dog | Not related |
| mRNA-1273 25 µg | Bronchial hyperreactivity | 96 days, Dose 2 | Hospitalized for 24 hour observation | Not related |
| mRNA-1273 25 µg | Seizure | 22 days, Dose 2 | None reported; Diagnosed with possible atypical seizure | Not related |
| Placebo | Rhinovirus infection<br>Asthma | 83 days, Dose 2<br>83 days, Dose 2 | Ongoing allergic rhinitis; attending daycare; family history of asthma; hospitalized on oxygen | Not related<br>Not related |
| Placebo | Abdominal wall abscess | 28 days, Dose 2 | Had recently undergone planned incisional hernia repair, orchidopexy, and bronchoscopy; Ongoing bronchopulmonary dysplasia; Chronic inhaled corticosteroid use | Not related |

i) SAE 6-23 months Pfizer[6]

| Treatment group | Preferred term | Time to Onset | Risk Factors, Pertinent Details | Assessment |
|---|---|---|---|---|
| BNT162b2 | RSV bronchiolitis | - | - | Non-related |
| BNT162b2 | RSV bronchiolitis | - | - | Non-related |
| BNT162b2 | RSV bronchiolitis | - | - | Non-related |
| BNT162b2 | RSV bronchiolitis | - | - | Non-related |
| BNT162b2 | RSV bronchiolitis | - | - | Non-related |
| BNT162b2 | Pneumonia | - | - | Non-related |
| BNT162b2 | Pneumonia | - | - | Non-related |
| BNT162b2 | Gastroenteritis | - | - | Non-related |
| BNT162b2 | Gastroenteritis | - | - | Non-related |
| BNT162b2 | Lower respiratory tract infection | - | - | Non-related |
| BNT162b2 | Lower respiratory tract infection | - | - | Non-related |
| BNT162b2 | Anaphylaxis | 14 days after dose 1 | Nut allergy | Non-related |
| BNT162b2 | Metapneumovirus infection, rhinovirus infection | - | - | Non-related |
| BNT162b2 | Anal abscess | - | - | Non-related |
| BNT162b2 | Accidental overdose | - | - | Non-related |
| BNT162b2 | Febrile convulsion | 38 days after dose 1 | Ear inflammation | Non-related |
| BNT162b2 | Seizure | 2 days after dose 2 | Respiratory tract infection | Non-related |
| Placebo | RSV bronchiolitis | - | - | Non-related |
| Placebo | RSV bronchiolitis | - | - | Non-related |
| Placebo | RSV bronchiolitis | - | - | Non-related |
| Placebo | Cyanosis | - | - | Non-related |
| Placebo | Cyanosis | - | - | Non-related |
| Placebo | Vomiting | - | - | Non-related |
| Placebo | anaphylaxis | - | - | Non-related |
| Placebo | norovirus gastroenteritis | - | - | Non-related |

| | | | | |
|---|---|---|---|---|
| Placebo | rotavirus gastroenteritis | - | - | Non-related |
| Placebo | tonsillitis | - | - | Non-related |
| Placebo | pneumomediastinum, respiratory distress | - | - | Non-related |
| Placebo | feeding intolerance, hypoglycemia, | - | - | Non-related |
| Placebo | head injury | - | - | Non-related |
| Placebo | second degree burns, thermal burns. | - | - | Non-related |

j) SAE 6-23 months Moderna[3,4]

| Treatment Group | SAE Preferred Term | Time to Onset after Most Recent Dose | Risk factors/pertinent details | Assessment |
|---|---|---|---|---|
| mRNA-1273 25 µg | Pyrexia<br>Febrile convulsion | Day of Dose 1<br>1 day after Dose 1 | Fever onset 6 hours post-vaccination; unwitnessed febrile convulsion 1 day after; rash developed 3 days after fever (see narrative above) | Related<br>Related |
| mRNA-1273 25 µg | Mastoiditis | 3 days after Dose 1 | Intermittent fevers 2 months prior to event onset; History of otitis media; Tested positive for adenovirus during hospitalization | Not related |
| mRNA-1273 25 µg | Metapneumovirus infection | 4 days after Dose 1 | Hospitalized in intensive care unit for 3 days | Not related |
| mRNA-1273 25 µg | Electrolyte imbalance | 8 days after Dose 1 | Hospitalized; concurrent RSV infection with respiratory distress and dehydration | Not related |
| mRNA-1273 25 µg | Rhinovirus infection | 8 days after Dose 1 | Hospitalized for 1 day. Reported by mother to have fever of 106o F. | Not related |
| mRNA-1273 25 µg | Foreign body in respiratory tract | 15 days after Dose 1 | Unspecified foreign body in respiratory tract, required bronchoscopy assisted removal | Not related |
| mRNA-1273 25 µg | Bronchiolitis | 17 days after Dose 1 | Food allergy (nuts), eczema; hospitalized on high-flow nasal cannula; viral panel negative | Not related |
| mRNA-1273 25 µg | Febrile convulsion | 21 days after Dose 1 | Medical history of intermittent fevers with rash; Fever for previous 2 days; Infectious disease and rheumatology diagnosed with PFAPA+; Received Dose 2 with no associated AEs | Not related |
| mRNA-1273 25 µg | Erythema multiforme | 35 days after Dose 1 | History of eczema and peanut allergy; exposed to peanut butter and amoxicillin (day 8 at onset of symptoms of EM) | Not related |
| mRNA-1273 25 µg | Adenovirus infection | 35 days after Dose 1 | Hospitalized due to concern for MIS-C vs Kawasaki Disease (KD). Found to be positive for adenovirus on PCR. SARS-CoV-2 negative; KD diagnosis excluded | Not related |
| mRNA-1273 25 µg | Asthma | 31 days after Dose 2 | Prodromal URI symptoms, temperature of 100.3°F, diagnosis of pneumonia at urgent care and started on amoxicillin; | Not related |

| | | | | |
|---|---|---|---|---|
| | | | hospitalized 4 days later for respiratory distress requiring high-flow nasal cannula, CXR at hospital: viral or reactive airway disease; viral panel negative | |
| mRNA-1273 25 µg | Diabetic ketoacidosis Type 1 diabetes mellitus | 37 days after Dose 2 | Family history of type 1 diabetes | Related |
| mRNA-1273 25 µg | Croup infectious | 43 days after Dose 2 | Inflammation of upper respiratory tract after recent adenoidectomy and turbinate reduction | Not related |
| mRNA-1273 25 µg | Gastroenteritis viral | 43 days after Dose 2 | Hospitalized (2 days) | Not related |
| mRNA-1273 25 µg | Febrile convulsion | 66 days after Dose 2 | Daycare reported possible seizure and tactile fever after a nap; fever 101.5 and normal exam in ED | Not related |
| Placebo | Bronchiolitis; Rhinovirus infection; Acute respiratory failure | 29 days after Dose 1 | Hospitalized on high flow nasal cannula | Not related |

**Supplementary Table 2**. Adverse events of special interest (AESI): The FDA defined list of AESI adapted from the Brighton Collaboration to be relevant to COVID-19 vaccines was used to assess cases in the study trials (supplementary table).

Both Pfizer and Moderna trials additionally described AEs of clinical interest regardless of the inclusion on the AESI list, these events were not included in our analysis.

### 9 Appendix A. Adverse Events of Special Interest

**Table 94. Adverse Events of Special Interest**

| Adverse Event | Additional Notes |
|---|---|
| Anosmia, ageusia | New onset COVID associated or idiopathic events without other etiology excluding congenital etiologies or trauma |
| Subacute thyroiditis | Including but not limited to events of atrophic thyroiditis, autoimmune thyroiditis, immune-mediated thyroiditis, silent thyroiditis, thyrotoxicosis and thyroiditis |
| Acute pancreatitis | Including but not limited to events of autoimmune pancreatitis, immune-mediated pancreatitis, ischemic pancreatitis, edematous pancreatitis, pancreatitis, acute pancreatitis, hemorrhagic pancreatitis, necrotizing pancreatitis, viral pancreatitis, and subacute pancreatitis<br>Excluding known etiologic causes of pancreatitis (alcohol, gallstones, trauma, recent invasive procedures) |
| Appendicitis | Include any event of appendicitis |
| Rhabdomyolysis | New onset rhabdomyolysis without known etiology such as excessive exercise or trauma |
| Acute respiratory distress syndrome (ARDS) | Including but not limited to new events of ARDS and respiratory failure. |
| Coagulation disorders | Including but not limited to thromboembolic and bleeding disorders, disseminated intravascular coagulation, pulmonary embolism, deep vein thrombosis |
| Acute cardiovascular injury | Including but not limited to myocarditis, pericarditis, microangiopathy, coronary artery disease, arrhythmia, stress cardiomyopathy, heart failure, or acute myocardial infarction |
| Acute kidney injury | Include events with idiopathic or autoimmune etiologies<br>Exclude events with clear alternate etiology (trauma, infection, tumor, or iatrogenic causes such as medications or radiocontrast, etc.)<br>Include all cases that meet the following criteria<br>Increase in serum creatinine by ≥ 0.3 mg/dl (≥ 26.5 µmol/L) within 48 hours;<br>OR<br>Increase in serum creatinine to ≥ 1.5 times baseline, known or presumed to have occurred within prior 7 days<br>OR<br>Urine volume ≤ 0.5 mL/kg/hour for 6 hours |
| Acute liver injury | Include events with idiopathic or autoimmune etiologies<br>Exclude events with clear alternate etiology (trauma, infection, tumor, etc.)<br>Include all cases that meet the following criteria<br>3-fold elevation above the upper normal limit for ALT or AST OR<br>>2-fold elevation above the upper normal limit for total serum bilirubin or GGT or ALP |
| Dermatologic findings | Chilblain-like lesions<br>Single organ cutaneous vasculitis<br>Erythema multiforme<br>Bullous rashes<br>Severe cutaneous adverse reactions including but not limited to: Stevens-Johnson Syndrome, Toxic Epidermal Necrolysis, Drug Reaction with Eosinophilia and Systemic Symptoms and fixed drug eruptions |

| Adverse Event | Additional Notes |
|---|---|
| Multisystem inflammatory disorders | Multisystem inflammatory syndrome in adults<br>Multisystem inflammatory syndrome in children<br>Kawasaki's disease |
| Thrombocytopenia | Platelet counts <150 × $10^9$<br>Including but not limited to immune thrombocytopenia, platelet production decreased, thrombocytopenia, thrombocytopenic purpura, thrombotic thrombocytopenic purpura, or HELLP syndrome |
| Acute aseptic arthritis | New onset aseptic arthritis without clear alternate etiology (e.g., gout, osteoarthritis, and trauma) |
| New onset of or worsening of neurologic disease | Including but not limited to:<br>Guillain-Barre syndrome<br>Acute disseminated encephalomyelitis<br>Peripheral facial nerve palsy (Bell's palsy)<br>Transverse myelitis<br>Encephalitis/encephalomyelitis<br>Aseptic meningitis<br>Febrile seizures<br>Generalized seizures/convulsions<br>Stroke (hemorrhagic and non-hemorrhagic)<br>Narcolepsy |
| Anaphylaxis | Anaphylaxis as defined per protocol.<br>Follow reporting procedures in protocol Section 7.4.5 |
| Other syndromes | Fibromyalgia<br>Postural orthostatic tachycardia syndrome<br>Chronic fatigue syndrome (includes myalgic encephalomyelitis and post viral fatigue syndrome)<br>Myasthenia gravis |

**Supplementary Table 2 a-j.** Description of AESIs in the included studies according to vaccine type and age group. Other relevant cases of interest not included as AESIs in study trials are described (if such were mentioned in the study reports). Assessment (related or non-related to vaccine during phase 3) by the trial study investigators are also presented.

a) AESI 12-15 years Pfizer[2]

| Treatment group | Preferred term | Time to Onset | Comment | Assessment |
|---|---|---|---|---|
| Placebo | Appendicitis | | | Not related |
| Placebo | Appendicitis | | | Not related |

b) AESI 12-17 years Moderna[3,4]

| Treatment group | Preferred term | Time to Onset | Previous | Assessment |
|---|---|---|---|---|
| mRNA-1273 | Hypersensitivity* | 11 days after dose 1 | - | Delayed cutaneous reaction deemed not an AESI |

* Not included in analysis

*From 8. November 2021-january 31, 2022, not included as no placebo group:*

*Adverse events of clinical interest*

*At the time of the May 8, 2021, data cutoff, the only AESI as specified by the study protocol was MIS-C. Since then, the protocol has been revised to include collection of AESIs based on the list of potential COVID-19 or COVID-19 vaccine related AESIs developed by the Brighton Collaboration (Appendix A). AEs reported in study P203 were retrospectively reviewed to identify AESIs. Through the January 31, 2022, data cutoff, among mRNA-1273 recipients, a total of 16 AESIs were reported in 13 participants (0.5%).*

*8 anosmia/ageusia (concurrent covid inf), 1 epilepsy, 1 aseptic meningitis, 1 hypersensitivity, 2 appendicitis.*

*In addition to the events noted above, there was one late-breaking event of chest pain reported after the January 31, 2022, data cutoff which was clinically concerning for vaccine-associated myocarditis.*

c) AESI 5-11 years Pfizer[5]

Initial + expansion group

| Treatment group | Preferred term | Time to Onset | Comment | Assessment |
|---|---|---|---|---|
| BNT162b2 | Tic | 7 days after dose 2 | | Related |
| BNT162b2 | Henoch-Schoenlein | 21 days after dose 1 | | Not related |
| BNT162b2 | Infectious arthritis | | SAE | Not related |

*Additional AEs of clinical interest, regardless of inclusion on the CDC AESI list, were evaluated based on sponsor safety data review. Not included in analysis.*

• Chest pain, non-cardiac chest pain, or chest discomfort were reported by 3 participants (0.2%) in the BNT162b2 group and 4 participants (0.5%) in the placebo group. None of these events had any reported cardiac involvement. Syncope was reported by 1 participant in the BNT162b2 group.

• One participant in the BNT162b2 group had a related AE of moderate paresthesia (bilateral lower extremity tingling) with onset at 1 day post-Dose 2 and reported as recovered/resolved 3 days after onset.

• One participant without a reported medical history in the BNT162b2 had a related AE Type IV hypersensitivity reaction characterized by a rash on the forehead, earlobe, and right forearm 3 days after Dose 1. A dermatologist diagnosed the rash as a Type IV hypersensitivity reaction and characterized the rash as ‘plaque, erythematous and minimal crusting’, and prescribed Triamcinolone and Benadryl creams. No prohibited concomitant treatments or nonstudy vaccines had been administered. The event was reported as resolving 18 days after onset without sequelae. This participant had no other reported AEs. This participant received Dose 2 without any additional AEs reported post-dose.

• One participant in the BNT162b2 group had a related AE of moderate angioedema reported as ‘perioral and periorbital angioedema due to allergic reaction’ and concurrent urticaria reported as ‘hives of the face and back caused due to allergic reaction’, both with onset of 2 days after Dose 2, and was reported as resolved 2 days after onset. Participant’s medical history included past allergy (hypersensitivity with mild rash) to a vaccine, Sever’s disease, contact dermatitis and seasonal allergies. This participant received no prohibited concomitant treatments or nonstudy vaccines. An analysis of angioedema cases reported in the BNT162b2 group included angioedema (see above) and urticaria (n=3). Two of the cases of urticaria were considered by the investigator as related to study intervention; one is described above (concurrent with angioedema).

• The second case of urticaria was mild (‘itchy’, ‘bilateral on hands and forearms’) with onset at 6 days after Dose 1 and resolved within 2 days, reported shortly after an AE of mild injection site erythema at 3 days after Dose 1, in a participant with no relevant medical history and no receipt of prohibited concomitant medications or nonstudy vaccines. This participant received Dose 2 without any reported post-dose AEs.

• Arthralgia was reported by a total of 2 participants, 1 each in the BNT162b2 (0.1%) and placebo (0.1%) groups.

d) AESI 6-11 years Moderna[3,4]

| Treatment group | Preferred term | Time to Onset | Previous | Assessment |
|---|---|---|---|---|
| mRNA-1273 | Chest pain and dyspnea* | 3 days after dose 2 | - | Related, but not a protocol-specified AESI |
| mRNA-1273 | Appendicitis | | | |
| mRNA-1273 | Appendicitis | | | |
| mRNA-1273 | Ageusia/anosmia | 25 days after dose 2 | Concurrent myalgia, diarrhea and injection site pain. No COVID. | |
| mRNA-1273 | Anosmia | | | |
| Placebo | Ageusia/anosmia | Occurring along with COVID-19 | | |
| Placebo | Anosmia | Occurring along with COVID-19 | | |

* Included as AESI in analysis

e) AESI 2-4 years Pfizer[6]

| Treatment group | Preferred term | Time to Onset | Comment | Assessment |
|---|---|---|---|---|
| BNT162b2 | Appendicitis | 105 days after dose 2 | | Not related |
| BNT162b2 | Appendicitis | 11 days after dose 2 | | Not related |
| BNT162b2 | Status epilepticus | 18 days after dose 1 | History of seizures. SAE | Not related |
| BNT162b2 | Epilepsy | 48 days after dose 2 | SAE | Not related |
| BNT162b2 | Febrile convulsions | 21 days after dose 1 | SAE | Not related |
| BNT162b2 | Febrile convulsions | 42 days after dose 2 | | Not related |
| Placebo | Convulsions | | | Not related |
| Placebo | Convulsions | | | Not related |
| Placebo | Convulsions | | | Not related |
| Placebo | Convulsions | | | Not related |

f) AESI 2-5 years Moderna[3,4]

| Treatment group | Preferred term | Time to Onset | Comment | Assessment |
|---|---|---|---|---|
| mRNA-1273 | Erythema multiforme | 3 days after dose 2 | - | Possibly related |
| mRNA-1273 | Erythema multiforme | 3 days after Dose 1 (in the setting of concomitant amoxicillin), 4 weeks after Dose 2, and 6 weeks after Dose 2 | - | Not related |
| mRNA-1273 | Chest pain | 5 days after dose 2 | - | Possibly related |
| mRNA-1273 | Food allergy | | - | Not related |
| mRNA-1273 | Seizure | 25 days after dose 2 | - | Not related |
| mRNA-1273 | Kawasaki Disease* | 76 days after dose 2 | Concurrent adenovirus and rhinovirus, negative for COVID-19 | Not related |
| Placebo | Henoch-Schönlein purpura | 3 days after dose 2 | Subsequent glycosuria and ageusia/anosmia with COVID-19 | Not related |
| Placebo | MIS-C | 113 days after dose 2 | Asymptomatic COVID-19 infections 37 days prior | Not related |

* After data cut-off, not included in analysis

g) AESI 6-23 months Pfizer[6]

| Treatment group | Preferred term | Time to Onset | Comment | Assessment |
|---|---|---|---|---|
| BNT162b2 | Convulsions | 3 days after dose 2 | Reported as SAE | Not related |
| BNT162b2 | Convulsions | 164 days after dose 2 | | Not related |
| BNT162b2 | Febrile convulsions | >30 days after vaccination | Reported as SAE | Not related |
| BNT162b2 | Febrile convulsions | >30 days after vaccination | | Not related |
| Placebo | Convulsions | | | Not related |

h) AESI 6-23 months Moderna[3,4]

| Treatment group | Preferred term | Time to Onset | Comment | Assessment |
|---|---|---|---|---|
| mRNA-1273 | Febrile convulsion | 1 day after dose 1 | - | Related |
| mRNA-1273 | Febrile convulsion | 21 days after dose 1 | Suspected PFAPA | Not related |
| mRNA-1273 | Liver injury | 2 days after dose 2 | Elevated alanine aminotransferase (ALT) and aspartate aminotransferase (AST) | Related |
| mRNA-1273 | Erythema multiforme | | - | Not related |
| Placebo | Acute respiratory failure, bronchiolitis, and rhinovirus infection | 29 days after dose 1 | - | Not related |

* Not included in AESI analysis as cases were after data cut-off.

**Supplementary Table 3. Cochrane Risk of Bias tool evaluating the included RCTs.**

| First author | Sequence generation | Allocation concealment | Blinding of participants and personnel | Blinding of outcome assessment | Incomplete outcome data | Selective reporting | Selective reporting supporting text |
|---|---|---|---|---|---|---|---|
| Frenck | Low | Low | Low | Low | Low | High | No uniformity in the reporting of follow-up in main article, supplementary material, and FDA and EMA reports presenting data from the same studies. |
| Walter | Low | Low | Low | Low | High | High | FDA and EMA reports on the same study describe an additional expansion group not included in main article, where more SAEs are reported than in the initial group. |
| Muñoz | Low | Low | Low | Low | Low | High | No uniformity in the reporting of follow-up in main article, supplementary material, and FDA and EMA reports presenting data from the same studies. |
| Ali | Low | Low | Low | Low | Low | High | Exact number of SAEs not reported in main text, and number of SAEs in supplementary lower than in EMA/FDA reports due to different follow-up time. |
| Creech | Low | Low | Low | Low | Low | High | No uniformity in the reporting of AEs and SAEs in main article, supplementary material, and FDA and EMA reports presenting data from the same studies due to different follow-up time |
| Anderson | Low | Low | Low | Low | Low | High | No uniformity in the reporting of AEs and SAEs in main article, supplementary material, and FDA and EMA reports presenting data from the same study due to different follow-up time. |

**Supplementary Table 4a. Health outcomes and relative risk calculations for children included in the Pfizer phase 3 trials (% (n)). Overall analyses stratified by both age and vaccine type. Trials with no information on event (marked with -) not included in overall RR calculations.**

Trial reports provided categories for all adverse events, defined using System Organ Class (SOC) groups. To investigate the two mRNA vaccine's effect on non-COVID-19 infections, we chose to include the total number of disease events within each of the following SOCs: "infections and infestations", "respiratory thoracic and mediastinal disorders", "gastrointestinal disorders", and "skin and subcutaneous tissue disorders". In addition, we also screened the tables for event numbers and chose to also include "nervous system disorders" and "musculoskeletal and connective tissue disorders", as there were both presenting enough cases to merit statistical analysis.[1]

| **Pfizer** | | | | | | | | | | |
|---|---|---|---|---|---|---|---|---|---|---|
| Age group | 12 to 15 years | | 5 to 11 years | | 2 to 4 years | | 6 to 23 months | | Overall | |
| | | | Safety + expansion group | | | | | | | |
| | BNT162b2 (N=1131) | Placebo (N=1129) | BNT162b2 (N=3109) | Placebo (N=1538) | BNT162b2 (N=1835) | Placebo (N=915) | BNT162b2 (N=1178) | Placebo (N=598) | BNT162b2 (N=7253) | Placebo (N=4180) |
| **Subjects with outcome in % (n)** | | | | | | | | | | |
| Serious AE overall | 0.6 (7) | 0.2 (2) | 0.1 (4) | 0.1 (1) | 0.7 (12) | 0.9 (8) | 1.4 (17) | 2.3 (14) | 0.6 (40) | 0.6 (25) |
| RR (95% CI) | 3.49 (0.73-16.78) | | 1.98 (0.22-17.69) | | 0.75 (0.31-1.82) | | 0.62 (0.31-1.24) | | 0.89 (0.55-1.45) | |
| Serious AE - non-accident | 0.6 (7) | 0.2 (2) | <0.1 (1) | <0.1 (1) | 0.7 (12) | 0.8 (7) | 1.4 (16) | 2.0 (12) | 0.5 (36) | 0.5 (22) |
| RR (95% CI) | 3.49 (0.73-16.78) | | 0.49 (0.03-7.90) | | 0.85 (0.34-2.16) | | 0.68 (0.32-1.42) | | 0.92 (0.55-1.55) | |
| Serious AE - infectious | <0.1 (1) | 0 (0) | <0.1 (1) | 0 (0) | 0.4 (8) | 0.5 (5) | 1.2 (14) | 1.2 (7) | 0.3 (24) | 0.3 (12) |
| RR (95% CI) | 7.38 (0.15-372)* | | 4.46 (0.07-287)* | | 0.80 (0.26-2.43) | | 1.02 (0.41-2.50) | | 1.03 (0.52-2.04) | |
| Severe AE | 0.8 (9) | 0.3 (3) | 0.2 (8) | 0.1 (1) | 0.5 (9) | 0.7 (6) | 1.0 (12) | 1.7 (10) | 0.5 (38) | 0.5 (20) |
| RR (95% CI) | 2.99 (0.81-11.03) | | 3.96 (0.50-31.61) | | 0.75 (0.27-2.09) | | 0.61 (0.26-1.40) | | 1.11 (0.65-1.88) | |
| AESI | 0 (0) | 0.2 (2) | 0.1 (3) | 0 (0) | 0.3 (6) | 0.4 (4) | 0.3 (4) | 0.2 (1) | 0.2 (13) | 0.2 (7) |
| RR (95% CI) | 0.14 (0.01-2.16)* | | 4.46 (0.40-49.4)* | | 0.75 (0.21-2.64) | | 2.03 (0.23-18.13) | | 1.00 (0.38-2.61) | |
| **System Organ Class (SOC) groups in % (n)** | | | | | | | | | | |
| GI disorders | 1.2 (14) | 0.3 (3) | 1.6 (37) | 1.7 (19) | 4.5 (83) | 5.8 (53) | 8.9 (105) | 8.7 (52) | 3.3 (239) | 3.0 (127) |
| RR (95% CI) | **4.66 (1.34-16.17)** | | 0.96 (0.56-1.67) | | 0.78 (0.56-1.09) | | 1.03 (0.75-1.41) | | 0.98 (0.80-1.20) | |
| Infections/infestations | 0.6 (7) | 0.7 (8) | 1.9 (42) | 2.0 (18) | 2.1 (38) | 2.4 (22) | 9.8 (116) | 9.7 (58) | 2.8 (203) | 2.5 (106) |
| RR (95% CI) | 0.87 (0.32-2.40) | | 1.15 (0.67-2.00) | | 0.86 (0.51-1.45) | | 1.02 (0.75-1.37) | | 1.00 (0.79-1.25) | |
| Musculosceletal disorders | 0.8 (9) | 0.7 (8) | 0.5 (13) | 0.7 (8) | 0.4 (7) | 0.4 (4) | 0.1 (1) | 0.2 (1) | 0.4 (30) | 0.5 (21) |
| RR (95% CI) | 1.12 (0.43-2.90) | | 0.80 (0.33-1.94) | | 0.87 (0.26-2.97) | | 0.51 (0.03-8.10) | | 0.90 (0.52-1.58) | |

| | | | | | | | | | | |
|---|---|---|---|---|---|---|---|---|---|---|
| Nervous system disorders | 1.1 (13) | 0.6 (7) | 0.7 (19) | 0.4 (7) | 0.4 (7) | 0.5 (5) | 1.2 (14) | 0.5 (3) | 0.7 (53) | 0.5 (22) |
| RR (95% CI) | 1.85 (0.74-4.63) | | 1.34 (0.57-3.19) | | 0.70 (0.22-2.19) | | 2.37 (0.68-8.21) | | 1.47 (0.89-2.41) | |
| Respiratory disorders | 0.2 (2) | 0.4 (4) | 1.4 (37) | 1.2 (19) | 3.9 (72) | 3.3 (30) | 5.0 (59) | 3.5 (21) | 2.2 (170) | 1.8 (74) |
| RR (95% CI) | 0.50 (0.09-2.72) | | 0.96 (0.56-1.67) | | 1.20 (0.79-1.82) | | 1.43 (0.88-2.32) | | 1.17 (0.90-1.54) | |
| Skin and tissue disorders | 0.6 (7) | 1.2 (13) | 1.4 (38) | 0.8 (10) | 1.1 (20) | 0.8 (7) | 3.0 (35) | 2.8 (17) | 1.3 (100) | 2.4 (47) |
| RR (95% CI) | 0.54 (0.22-1.34) | | 1.88 (0.94-3.76) | | 1.42 (0.60-3.36) | | 1.05 (0.59-1.85) | | 1.18 (0.83-1.69) | |

Significant results in bold. Trials with no information on event (marked with -) or zero events in both vaccine and placebo groups not included in overall RR calculations. RRs were calculated as Mantel-Haenszel fixed-effect estimates stratified by vaccine in age groups and also stratified by age group in combined estimates (overall). *Peto OR.

**Supplementary Table 4b. Health outcomes and relative risk calculations for children included in the Moderna phase 3 trials. Overall analyses stratified by both age and vaccine type. Trials with no information on event (marked with -) not included in overall RR calculations.**

Trial reports provided categories for all adverse events, defined using System Organ Class (SOC) groups. To investigate the two mRNA vaccine's effect on non-COVID-19 infections, we chose to include the total number of disease events within each of the following SOCs: "infections and infestations", "respiratory thoracic and mediastinal disorders", "gastrointestinal disorders", and "skin and subcutaneous tissue disorders". In addition, we also screened the tables for event numbers and chose to also include "nervous system disorders" and "musculoskeletal and connective tissue disorders", as there were both presenting enough cases to merit statistical analysis.[1]

| **Moderna** | | | | | | | | | | |
|---|---|---|---|---|---|---|---|---|---|---|
| | 12 to 17 years | | 6 to 11 years | | 2 to 5 years | | 6 to 23 months | | Overall | |
| | mRNA-1273 (N=2486) | Placebo (N=1240) | mRNA-1273 (N=3007) | Placebo (N=995) | mRNA-1273 (N=3031) | Placebo (N=1007) | mRNA-1273 (N=1761) | Placebo (N=589) | mRNA-1273 (N=10285) | Placebo (N=3831) |
| **Subjects with outcome in % (n)** | | | | | | | | | | |
| Serious AE overall | 0.2 (6) 16[a] | 0.2 (2) | 0.2 (6) 11[b] | 0.2 (2) 0[c] | 0.3 (9) | 0.2 (2) | 0.9 (15) | 0.2 (1) | 0.4 (36) | 0.2 (7) |
| RR (95% CI) | 1.50 (0.30-7.40) | | 0.99 (0.20-4.91) | | 1.50 (0.32-6.91) | | 5.02 (0.66-37.90) | | 1.87 (0.83-4.21) | |
| Serious AE - non-accident | 0.2 (4) | 0.2 (2) | 0.2 (6) | 0.2 (2) | 0.3 (8) | 0.2 (2) | 0.8 (14) | 0.2 (1) | 0.3 (32) | 0.2 (7) |
| RR (95% CI) | 1.00 (0.18-5.44) | | 0.99 (0.20-4.91) | | 1.33 (0.28-6.25) | | 4.68 (0.62-35.53) | | 1.64 (0.72-3.73) | |
| Serious AE - infectious | 0 (0) | 0 (0) | 0.1 (3) | 0 .1 (1) | 0.2 (7) | 0.1 (1) | 0.6 (11) | 0.2 (1) | 0.2 (21) | <0.1 (3) |
| RR (95% CI) | N/A | | 0.99 (0.10-9.53) | | 2.33 (0.29-18.88) | | 3.68 (0.48-28.44) | | 2.33 (0.70-7.81) | |
| Severe AE | 0.4 (11) | 0.1 (1) | 0.4 (11) | 0.1 (1) | 0.7 (21) | 0.9 (9) | 1.0 (18) | 0.7 (4) | 0.6 (61) | 0.4 (15) |
| RR (95% CI) | 5.49 (0.71-42.45) | | 3.64 (0.47-28.16) | | 0.78 (0.36-1.69) | | 1.51 (0.51-4.43) | | 1.45 (0.83-2.52) | |
| AESI | 0 (0) | 0 (0) | 0.2 (5) | 0.2 (2) | 0.2 (5) | 0.2 (2) | 0.2 (4) | 0.2 (1) | 0.1 (14) | 0.1 (5) |
| RR (95% CI) | N/A | | 0.83 (0.16-4.26) | | 0.83 (0.16-4.27) | | 1.34 (0.15-11.95) | | 0.93 (0.34-2.58) | |
| **System Organ Class (SOC) groups in % (n)** | | | | | | | | | | |
| GI disorders | 1.1 (28) | 1.6 (20) | 2.7 (81) | 2.9 (29) | 4.4 (133) | 4.7 (47) | 9.7 (170) | 10.2 (60) | 4.0 (412) | 4.1 (156) |
| RR (95% CI) | 0.70 (0.40-1.23) | | 0.92 (0.61-1.41) | | 0.94 (0.68-1.30) | | 0.95 (0.72-1.25) | | 0.91 (0.76-1.09) | |
| Infections/infestations | 2.9 (71) | 3.1 (38) | 9.0 (272) | 7.3 (73) | 19.2 (582) | 17.6 (177) | 26.3 (464) | 26.1 (154) | 13.5 (1389) | 11.5 (442) |
| RR (95% CI) | 0.93 (0.63-1.37) | | 1.23 (0.96-1.58) | | 1.09 (0.94-1.27) | | 1.01 (0.86-1.18) | | 1.07 (0.97-1.18) | |
| Musculosceletal disorders | 2.3 (58) | 2.6 (32) | 1.6 (48) | 1.8 (18) | - | - | - | - | 1.9 (106) | 2.2 (50) |
| RR (95% CI) | 0.90 (0.59-1.38) | | 0.88 (0.52-1.51) | | - | | - | | 0.90 (0.64-1.25) | |
| Nervous system disorders | 2.7 (68) | 2.5 (31) | 2.8 (84) | 3.3 (33) | 1.1 (32) | 1.4 (14) | 2.2 (38) | 2.5 (15) | 2.2 (222) | 2.4 (93) |

| | | | | | | | | | | |
|---|---|---|---|---|---|---|---|---|---|---|
| RR (95% CI) | 1.09 (0.72-1.66) | | 0.84 (0.57-1.25) | | 0.76 (0.41-1.42) | | 0.85 (0.47-1.53) | | 0.91 (0.71-1.15) | |
| Respiratory disorders | 1.4 (34) | 1.0 (12) | 6.1 (184) | 7.0 (70) | 7.7 (233) | 8.3 (84) | 8.1 (143) | 7.8 (46) | 5.6 (594) | 5.5 (212) |
| RR (95% CI) | 1.41 (0.73-2.72) | | 0.87 (0.67-1.13) | | 0.92 (0.73-1.17) | | 1.04 (0.76-1.43) | | 0.95 (0.82-1.11) | |
| Skin and tissue disorders | 1.1 (28) | 0.6 (7) | 2.4 (71) | 1.0 (10) | 2.0 (62) | 1.4 (14) | 3.0 (52) | 3.6 (21) | 2.1 (213) | 1.4 (52) |
| RR (95% CI) | 2.00 (0.87-4.55) | | **2.35 (1.22-4.54)** | | 1.47 (0.83-2.62) | | 0.83 (0.50-1.36) | | **1.44 (1.07-1.95)** | |

[a]15 additional SAEs described in extended follow-up with no information on placebo group. Not included in SAE analysis.

[b,c]11 additional SAEs described in extended follow-up, including both blinded data and events in children from placebo group being unblinded and moved to vaccine group. No description on children in placebo group not crossing over, hence data not included in SAE analysis.

Significant results in bold. Trials with no information on event (marked with -) or zero events in both vaccine and placebo groups not included in overall RR calculations. RRs were calculated as Mantel-Haenszel fixed-effect estimates stratified by vaccine in age groups and also stratified by age group in combined estimates (overall). *Peto OR.

**Supplementary Table 5. Number needed to treat or harm for COVID-19 as well as primary and secondary outcomes by age group for Pfizer and Moderna vaccines combined.**

| | 12-15(17) years | 5(6)-11 years | 2-4(5) years | 6-23 months | Overall |
|---|---|---|---|---|---|
| **Pfizer + Moderna** | | | | | |
| COVID-19 | 115 (108 to 139) | 93 (84 to 123) | 40 (31 to 64) | 60 (39 to 370) | 72 (63 to 88) |
| SAE overall | -440 (NS) | -2817 (NS) | 2141 (NS) | 1592 (NS) | -2091 (NS) |
| Severe AE | -215 (-2470 to -58) | -455 (NS) | 559 (NS) | 775 (NS) | -883 (NS) |
| URT | - | -196 (NS) | 246 (NS) | 366 (NS) | -2560 (NS) |
| RSV | - | - (*) | -726 (NS) | -382 (NS) | -635 (-12544 to -287) |
| LRTI | - | - (*) | -278 (-96151 to -52) | -276 (NS) | -217 (-1217 to -79) |

A positive NNT indicates a beneficial effect (NNTB), corresponding to a RR <1 for the given event, whereas a negative value indicates a negative effect (NNTH), corresponding to a RR >1 for the given event. 95% CIs have been provided for statistically significant estimates, but not for non-significant (NS) estimates.
(*) No events in the control group.
